# Heterozygous germline *MSH3* mutations, and probably *MLH3* mutations, act as classical tumour suppressors, leading to excess somatic deletion mutations, signature ID4 and increased colorectal cancer risk

**DOI:** 10.64898/2026.07.22.26358679

**Authors:** Ignacio Soriano, Kitty Sherwood, Joseph C. Ward, Juan Fernandez-Tajes, Anusha Aman, Steve Thorn, Guler Gul, James Wilson, Anders Isaksson, Bengt Glimelius, Tobias Sjoblom, Claire Palles, Ian Tomlinson

## Abstract

**Background:** *MSH3* and *MLH3* are non-canonical DNA mismatch repair genes, involved in repairing insertion-deletion mutations. Colorectal cancer (CRC) and adenomas have been reported in patients with bi-allelic germline *MSH3* mutations, and in a very few bi-allelic *MLH3* mutation carriers.

**Objectives:** We hypothesised that germline loss-of-function *MSH3* and *MLH3* mutations were akin to constitutional mismatch repair deficiency (cMMRd) and Lynch syndrome, such that CRC could result from either bi-allelic germline mutations, or heterozygous germline mutations after second hits.

**Design:** Nearly 12,000 CRC and multiple polyp cases and 460,000 controls were studied. 2,023 patients underwent cancer genome sequencing.

**Results:** One CRC/multiple polyp case had bi-allelic *MSH3* mutations and another, bi-allelic *MLH3* mutations. *MSH3* and *MLH3* germline heterozygotes had an increased risk of CRC (respectively 2.2-fold, *P=*6.6x10^-5^ and 1.6-fold, *P=*0.028), owing to somatic ‘second hits’ that inactivated the wildtype allele. Single second hits sometimes inactivated both *MSH3* and the nearby *APC*gene. All CRCs with MSH3 or MLH3 deficiency were microsatellite-stable, but hypermutant. *D*eletions of *≥*2bp were particularly increased (∼12-fold) and signature ID4 was usually present (*P<*0.0001). CRCs from heterozygotes without ‘second hits’ showed no hypermutation.

**Conclusion:** The phenotypes of bi-allelic *MSH3* and *MLH3* mutation carriers resemble some cMMRd patients. Heterozygous germline *MSH3* and *MLH3* alleles have incomplete penetrance, but increase CRC risk *via* hypermutation, phenotypically resembling *PMS2-*mutant Lynch syndrome. A causal association with specific mutations has not previously been reported for ID4 in human tumours. ID4 probably does not have a single aetiology, but can result from MSH3 or MLH3 deficiency.

**Summary Box:** *What is already known on this topic – summarise the state of scientific knowledge on this subject before you did your study and why this study needed to be done.:* Lynch syndrome and constitutional mismatch repair deficiency (cMMRd) are established cancer syndromes, respectively resulting from mono-allelic (heterozygous) and bi-allelic germline mismatch repair (MMR) mutations. Germline bi-allelic *MSH3* mutations are known to predispose to a recessive syndrome of multiple colorectal adenomas and colorectal carcinoma (CRC). Weaker evidence, supported herein, suggests that bi-allelic *MLH3* mutations cause a similar syndrome. Occasional, relatively small studies have reported CRC cases with heterozygous germline loss-of-function *MSH3* or *MLH3* mutations, leading to suggestions that these mutations increase CRC risk. However, these studies have essentially been anecdotal, presenting no statistical evidence of increased cancer risk or an identified causal mechanism. MSH3 and MLH3 deficiency *in vitro* have been linked to insertion-deletion mutations of a few bases, but evidence from human tumours is lacking. We hypothesised that *MSH3* and/or *MLH3* resemble Lynch and cMMRd genes, in that they predispose to CRC (and possibly other cancers) when heterozygous, as well as in the much rarer bi-allelic mutant state.

*What this study adds – summarise what we now know as a result of this study that we did not know before.:* In a large case-control analysis, we have shown that heterozygous loss-of-function (LoF) germline *MSH3* mutations increase CRC risk about 2.2-fold. Like the Lynch syndrome genes, *MSH3* in this context acts as a tumour suppressor (TSG), with somatic second hits in cancers. The second hits are sometimes loss-of-heterozygosity changes, and, of note, one LOH event can sometimes inactivate both *MSH3* (chr5q14.1) and the major CRC driver gene *APC*(chr5q22.2). Heterozygous germline *MLH3* mutations also appear to increase CRC risk (1.6-fold) in a similar fashion. The relative risks in heterozygotes are of a size that *MSH3* and *MLH3* could be said to be moderate risk CRC genes, among the first of this class to be found. Mismatch repair deficiency by loss of MSH3 or MLH3 function does not generally cause MSI, but results in relatively mild hypermutation *via* deletions of *≥*2bp, 1bp deletions at oligonucleotide tracts, and mutational signature ID4. ID4 has no previously established cause in human tumours, but has been associated with *Rnaseh2b;Tp53*deficiency in mouse and cell models. MSH3-deficient CRCs otherwise largely resemble MSS CRCs.

*How this study might affect research, practice or policy – summarise the implications of this study.:* Given that Lynch and cMMRd include patients with germline mutations in genes like *PMS2* with weaker effects on cancer risk, we suggest that *MSH3* and *MLH3* could also be regarded as genes for these syndromes. The two genes would reasonably be included in panels used to investigate the causes of CRC and/or multiple polyps in clinical practice. The size of the increased risk suggests that heterozygous gene carriers could be offered modestly enhanced surveillance for CRC (e.g. by earlier or more frequent faecal immunochemical testing). Research into genetic variants in other DNA repair genes, even if not historically associated with CRC risk, may prove fruitful, especially if based on large genome sequencing studies of both germline and tumour genotypes.

## Introduction

Mono-allelic germline mutations in the DNA mismatch repair (MMR) genes *MSH2, MLH1, MSH6* and *PMS2* predispose to Lynch syndrome [1]. The phenotypes and cancer risks associated with each gene vary, but the principal features are early-onset colorectal and endometrial cancers, with smaller increased risks of malignancies of other tissues and organs. Following somatic inactivation of the germline wildtype allele, Lynch syndrome tumours are hypermutant and are MMR-deficient (MMRd). The resulting ‘classical’ MMR-deficiency manifests in cancers as microsatellite instability (MSI), owing to single- or double-base insertion-deletion (indel) mutations in short tandem repeat (STR) DNA sequences. MSI sometimes results in loss-of-function (LoF) mutations in driver genes such as *TGFBR2* and *B2M*that contain protein-coding STRs. After Lynch syndrome was characterised, bi-allelic germline MMR mutations were found to cause constitutional mismatch repair deficiency (cMMRd) [2, 3], a syndrome typified by café-au-lait spots, brain tumours and other childhood malignancies, and multiple bowel polyps [4, 5].

Bi-allelic LoF germline mutations in the ‘minor’ (or ‘non-canonical’) MMR gene *MSH3* have been found in about 10 kindreds [6, 7, 8], most of whom have multiple colorectal adenomas and colorectal carcinoma (CRC), but no clear excess of extra-colonic tumours. Inheritance appears to be recessive, with relatively high penetrance. A small number of these individuals’ tumours have been analysed for somatic mutations, but no clear pathogenic mechanism downstream of MSH3 deficiency has been identified [6, 9]. Bi-allelic germline mutations in the related gene, *MLH3*, have also been reported in two Scandinavian multiple adenoma families, one with a founder mutation from Sweden and Finland [10] and one from Denmark [11]. However, another report of a similar patient cast doubt on the pathogenicity of *MLH3* mutations in this context [12].

MSH3-MSH2 and MLH3-MLH1 heterodimers may act in the MutS*β* and MutL*γ* complexes respectively to repair indels of a few base pairs. However, colorectal polyps from MLH3-deficient patients have shown no instability at mono-, di-, tri-, or tetranucleotide repeats [10]. Overall, neither *MSH3* nor *MLH3* is routinely tested in clinical practice, and a molecular mechanism of tumorigenesis has not been established. We hypothesised that germline loss-of-function *MSH3* and *MLH3* mutations were akin to constitutional mismatch repair deficiency and Lynch syndrome, such that either bi-allelic mutations, or heterozygous mutations accompanied by somatic second hits, could cause CRC. We tested this model in large sets of CRC patients, including some whose cancers had undergone whole genome sequencing (WGS).

## Results

### LoF-of-function germline variants in CRC patients

We searched for pathogenic (loss-of-function) germline *MSH3* and *MLH3* variants in 3,135 unrelated UK patients with CRC and/or multiple polyps who had undergone WGS as part of the UK 100,000 Genomes project (100kGP) or the CORGI study (**Methods**) [13, 14]. Bi-allelic germline *MSH3* or *MLH3* variants were present in two 100kGP cases, both of whom had multiple adenomatous polyps and CRC (**Table 1**; **Figure 1; Supplementary Tables 1 & 2**). One of these patients was recruited on the basis of their multiple polyp phenotype, but the other was enrolled into the sporadic cancer arm of the 100kGP, for which most CRC cases presenting to a major general hospital in England were eligible.

**Figure 1.**
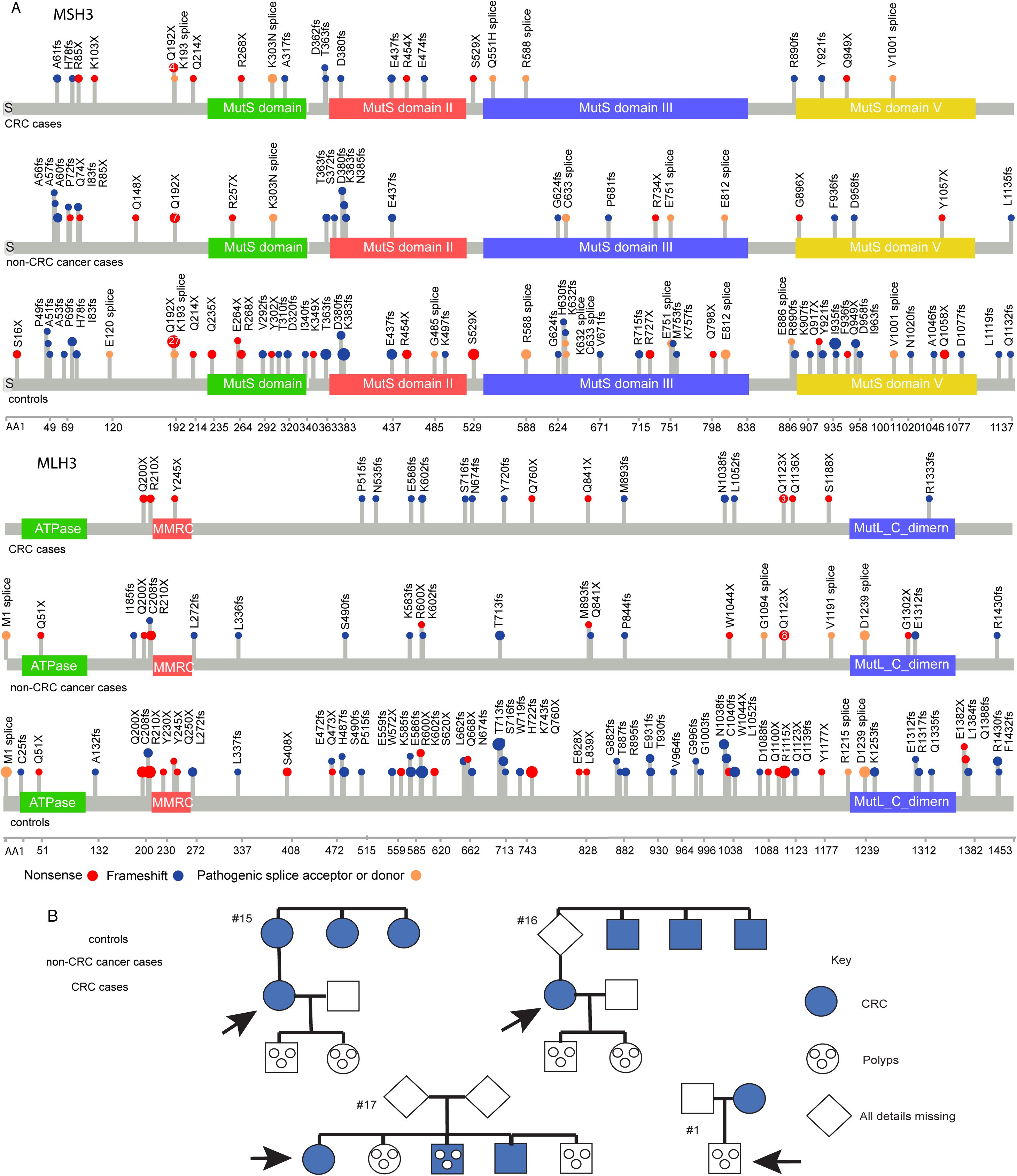
Germline *MSH3* and *MLH3* LoF mutations and family pedigrees. (A) Lollipop diagrams of putative LoF MSH3 and MLH3 germline mutations in CRC cases, extracolonic cancer cases and controls from 100kGP, CORGI and UKB. For MSH3, germline mutations are shown for 31 CRC cases, 46 other cancer cases and 186 controls; corresponding numbers for *MLH3* are respectively 24, 47 and 259. Median ages (range) were 61 (40-70) for CRC cases, 57 (41-69) for other cancer cases, and 55 (39-72) for controls. Lollipop sticks show the locations of putative pathogenic (LoF) germline variants (amino acid position for each gene shown below the plots). Head colours show type of mutation (red=nonsense, blue=frameshift, orange=splice), and head sizes are linearly proportional to the frequency of the mutation in each of the six data sets. The number of individuals with the most frequent mutation in each data set is shown within the lollipop head. Thus in the *MSH3* controls data, the most frequent mutation was Q192X, present in 27 individuals, whereas mutations such as E120splice, Q214X, E264X, Y302X, I340fs, G485, G624fs, V671fs, M753fs, Q798X, E886, F936fs, N1020fs and Q1132fs were each present in a single individual, and other mutations were at frequencies intermediate between these. There were no clear mutational hotspots in either gene. Our focus on protein-inactivating (LoF) mutations was pre-specified, but we also performed exploratory checks for germline missense mutations at highly conserved sites of predicted functional importance (e.g. ATP binding or phosphorylation targets). However, only two such variants were found and each had uncertain predicted pathogenicity; these individuals were therefore not included in our analyses. Plots were generated with SRPlot (http://www.bioinformatics.com.cn/plot_basic_lollipop_mutation_diagram_090_en). *(B) Simplified pedigrees of probands (arrowed) with a mono-allelic germline MSH3 or MLH3 mutation*. Pedigree numbering corresponds to **Table 1** and **Supplementary Tables 1 & 2**. Blue=CRC, circles=polyp(s), diamond=details missing. These cases were derived from the 100kGP multiple polyp domain (#1) or the CORGI study which is enriched for familial CRC (#s 15, 16 and 17). Note that neither tumour sequencing nor mutation co-segregation data were available for these CORGI individuals.

**Table 1.**
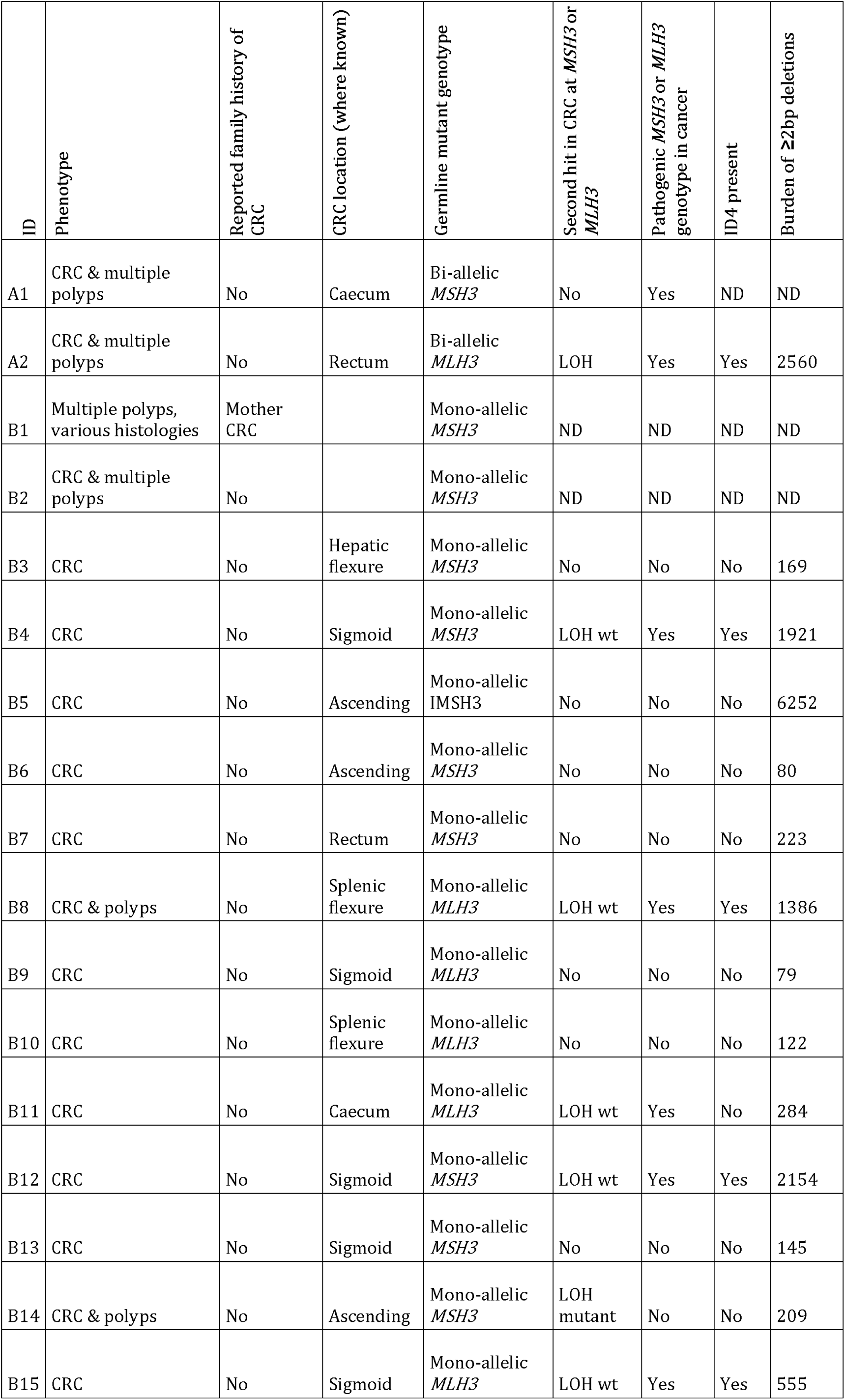
Clinical and molecular data from 100kGP CRC cases with pathogenic germline MSH3 or MLH3 variants. Full data from these individuals and others included in the study are available in **Supplementary Tables 1 & 2**. Polyps were coded as adenomatous or hyperplastic, or not specified, but sizes, numbers and detailed morphology were variably available. Ages are shown by decade to preserve confidentiality. Family history was not generally reported for 100kGP cancer domain cases. Full histological details and locations of cancers were not always available. ID4 metrics are shown for cancers with and without somatic loss of MSH3 or MLH3 function. Patient A1 had prior chemotherapy, and hence was not evaluated for mutational signatures as part of this project. The allelic loss in patient A2’s CRC had no predicted additional functional consequences. For cancers B4 and B12, LOH events at *APC*and *MSH3* were concordant, supporting a model in which a single event caused loss of the wildtype alleles at both loci. Case B15 was not included in the case-control (association) analyses of CRC: whilst we strongly suspected a germline *MLH3* mutation in this patient, this could not be established with certainty (further details in Figure 2). wt=wildtype allele.

Seventeen 100kGP/CORGI CRC patients carried heterozygous (mono-allelic), germline LoF *MSH3* (n=12) or *MLH3* (n=5) variants (**Table 1; Supplementary Tables 1 & 2**). Although polyps and family history were variably reported, four of the heterozygotes’ kindreds were established as having CRC in multiple generations (**Figure 1B**). The CRCs of 15 of the 17 patients had undergone genome sequencing. After excluding a patient who had previously received systemic genotoxic chemotherapy (A-1, **Table 1; Supplementary Table 1**), five (one germline bi-allelic *MLH3,* two *MSH3* with LOH, two *MLH3* with LOH) of these tumours had second hits inactivating the wildtype allele owing to loss of heterozygosity (LOH). Bi-allelic *MSH3* or *MLH3* mutations were thus created in these tumours, in classical tumour suppressor (TSG) fashion (**Table 1; Supplementary Table 1; Figure 2**). For *MSH3,* both of the detected second hits appeared to be ‘serendipitous’, since they appeared primarily to target the major polyp and CRC driver TSG *APC*, which is about 33Mb distal to *MSH3* on the long arm of chromosome 5 (**Figure 2A**). As was the case here, *APC*LOH events frequently have a proximal breakpoint between 65Mb and 75Mb LOH [15] and these events can also cause LOH of a germline *MSH3* mutation (chr5:80.7-80.9Mb, GRCh38/hg38) *in cis* with the *APC*first hit. The two second hit LOH events involving MLH3 in our CRCs appeared to be isolated changes, not targeting any nearby CRC driver gene such as AKT1. Whilst 14q LOH and 5q LOH events have similar frequencies, 31% and 38% respectively in MSS CRCs [16], the former tend to occur in advanced malignant lesions [17], whereas the latter classically cause adenoma initiation. We hypothesise that this could allow MSH3 to have a greater influence than MLH3 on colorectal carcinogenesis, including the inactivation of TSGs (e.g. SMAD4 and TP53) subsequent to APC but prior to malignant progression [18] (**Table 1; Supplementary Table 1**).

**Figure 2.**
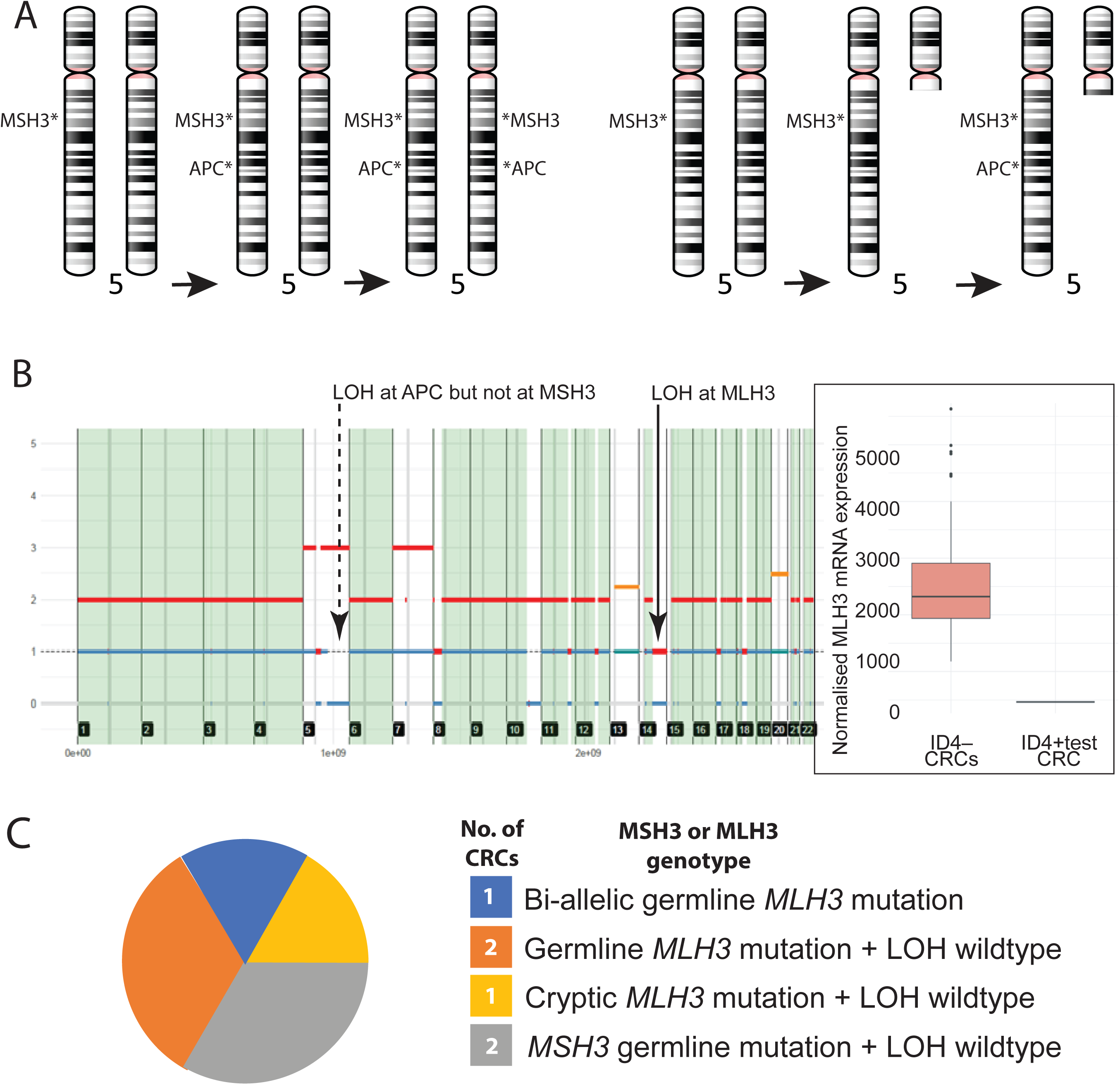
Mechanisms of MSH3/MLH3 deficiency. *(A) Chromosome 5 ideograms showing mechanism of MSH3 and APC inactivation by a single LOH event in carriers of germline mono-allelic MSH3 mutations*. In our preferred model (left), there is a first somatic *APC*mutation *in cis*with the germline *MSH3* mutation, followed by a single LOH event (which is usually copy-neutral at *APC*) that removes the wildtype copies of both genes, thus ‘serendipitously’ inactivating both *APC*and *MSH3*, leading to a higher risk of CRC. An alternative model (right) is also possible, in which an initial large (gene-level or greater) LOH event by somatic deletion is the first somatic mutation, removing the wildtype copy of *MSH3* and also one copy of *APC*, prior to a second hit at *APC* occurring on the remaining long arm of chr5q. We believe the latter model to be less likely model because *APC*LOH events (including those in the *MSH3* mutation carriers) usually occur by copy-neutral mechanisms, which is only effective if there is a pre-existing *APC* mutation on the other chromosome. A single LOH event involving *MSH3* and *APC* occurred in the CRCs of 100kGP cases B4 and B12 (**Supplementary Table 1**). LOH events that involve *APC* alone and do not extend to *MSH3* (e.g. CRC B3, **Supplementary Table 1**), and somatic nonsense or frameshift LoF *APC* mutations (**Supplementary Tables 1 & 2**), evidently have no effect on the wildtype *MSH3* allele and any tumour that grows in that cell may proceed along the canonical molecular pathways of colorectal tumorigenesis, despite the germline *MSH3* variant. *(B) Analysis of the ID4-positive CRC without an identified germline mutation. The main plot shows copy number analysis from chromosome 1 to 22 in this CRC, showing segmental dosage (red total copy number, blue minor haplotype copy number)*. The main plot shows copy number analysis from chromosome 1 to 22 in this CRC, showing segmental dosage (red total copy number, blue minor haplotype copy number). This CRC had ID4, but no obvious cause of bi-allelic *MSH3* or *MLH3* inactivation, including pathogenic non-coding variants, structural variants and promoter methylation (details not shown). The cancer did, however, show somatic LOH by copy loss LOH at *MLH3* (chr5:51.2-106.9Mb, solid line and arrow), with retention of a single copy of *MLH3*. *MSH3* (chr5:80Mb) showed no evidence of inactivation, with total copy number of 4 and minor copy number of 1. RNAseq data for this cancer (“ID4+ test CRC”) are shown in the box plot (centre line shows median, box limits upper and lower quartiles, and whiskers 1.5x inter-quartile range). There was exceptionally low *MLH3* mRNA expression, far below any of 231 other CRCs profiled (left box). Overall mRNA expression levels in the ID4+ cancer, specifically including *MSH3*, were unremarkable. The presence of ID4 in this cancer is thus explained by very low *MLH3* mRNA expression, which appears to result from somatic loss of one allele and a cryptic defect, which we presumed to be of germline origin, in the other. *(C) Breakdown of the putative origins of the MSH3/MLH3 deficiency in the six CRCs*. These patients were A-2, B-4, B-8, B-11, B-12 and B-15 from **Table 1** and **Supplementary Table 1**.

We subsequently identified a sixth CRC with loss of MLH3. This cancer did not harbour detectable LoF germline *MSH3* or *MLH3* mutations, but showed somatic LOH by copy loss at *MLH3* (chr5:51.2-106.9Mb, solid line and arrow), resulting in a single copy of *MLH3*. No pathogenic non-coding variants, structural variants and promoter methylation were found in this patient or their cancer (details not shown). However, RNAseq data were fortunately available. These data showed near-complete loss of *MLH3* mRNA expression in this cancer, at levels far below any of 231 other CRCs profiled with RNAseq (**Table 1**; **Figure 2B; Supplementary Table 1**), whilst overall mRNA expression levels, including *MSH3,* were unremarkable. The MSH3/MLH3-deficient status of this cancer was thus explained by its very low *MLH3* mRNA expression, which we surmise to have resulted from somatic loss of one allele and a cryptic defect, which we presumed to be of germline origin, on the remaining chromosome 14.

### Genomic features of MSH3/MLH3-deficient colorectal cancer

We analysed the set of six available CRCs with *MSH3/MLH3 deficiency*(**Figure 2C**), that is, bi-allelic LoF mutations of germline or somatic origin. All these cancers were MSI-negative according to msiNGS analysis [19] and their single base substitution (SBS) burdens, spectra and COSMIC mutational signatures were very similar to those of the microsatellite-stable/polymerase proofreading-proficient CRCs (termed MSS herein, n=1,619) that comprised the bulk of the available tumour WGS data (**Table 2**; **Figure 3A,B; Supplementary Table 3**). There were no secondary somatic mutations of other MMR genes in the MSH3/MLH3-deficient cancers, and none of the single base substitution (SBS) signatures (SBS6, SBS14, SBS15, SBS20, SBS26 and SBS44) specific to MMRd-MSI+ cancers was present. The SBS burdens of MSH3/MLH3-deficient cancers were commensurately lower than those of the 334 MSI+ tumours. However, the indel mutation profile of the MSH3/MLH3-deficient cancers was distinct from both the MSS and MSI+ CRCs (**Figure 3B,C; Supplementary Table 3**). The indel burdens of MSH3/MLH3-deficient cancers (median=4,200) were dwarfed by the number of indels in MSI+ tumours (median ∼142,000), which mostly comprised 1bp deletions in poly(T) tracts (median ∼100,000 per MSI+ CRC), and overall, MSI+ CRCs had a significant excess burden of most indel types over MSH3/MLH3-deficient cancers. However, a notable exception was several of the 83 indel channels that comprised *≥*2bp indels at STRs or sites of microhomology, for which burdens were similar in MSI+ and MSH3/MLH3-deficient cancers (**Supplementary Table 3**).

**Figure 3.** Mutational burdens and signatures in MSH3/MLH3-deficient cancers compared with MSI+ and MSS CRCs. *(A) Presence and activities (proportional burdens) of SBS, DBS and ID signatures detected in the six MSH3/MLH3-deficient CRCs*. Cancers are shown according to the IDs in **Table 1** and **Supplementary Table 1**. The only detected association between any signature and MSH3/MLH3- deficiency, compared with the bulk of MSS cancers, was the presence of ID4. *(B) Burdens (total numbers per cancer) of the four major somatic mutation types: single base substitutions (SBS); indels; somatic copy number alterations at arm level (CNAs); and structural variants (SVs) in the six MSH3/MLH3-deficient CRCs and in comparator groups (Lynch, all MSI+, MSS)*. ** P<0.01, *** P<0.001, **** P<0.0001 from Wilcoxon tests, comparing each group pairwise with the ID4-positive group. *(C) Activities (burdens relative to all indels) of specific types of indel in the six ID4-positive CRCs (orange) and in comparator groups (all MSI+ in purple and MSS in pink)*. The six MSH3/MLH3-deficient CRCs were compared pairwise against the other two groups using multiple logistic regression with robust variances, including age, sex, tumour purity and CRC location (proximal or distal colorectum) as co-variables. * P<0.05, ** P<0.01, *** P<0.001, **** P<0.0001. Further details are in **Table 2** and **Supplementary Tables 1 & 3**.

**Table 2.**
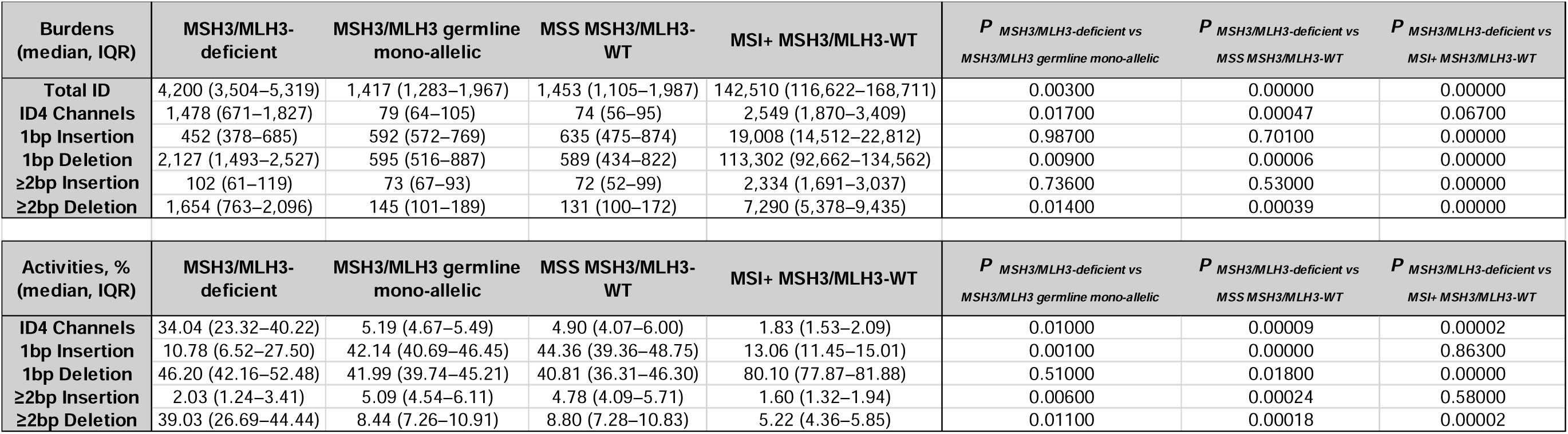
Summary of the burdens and activities of insertion-deletion mutations in colorectal cancers according to MSH3/MLH3 and MSI+ status. Medians and inter-quartile ranges are shown. Activity was measured as burden relative to the total indel burden in each cancer. MSH3/MLH3-Deficient cancers (n=6) had at least one germline loss-of-function MSH3 or MLH3 mutation and a bi-allelic mutant genotype in the cancer (e.g. owing to loss-of-heterozygosity). MSH3/MLH3 germline mono-allelic cancers (n=8) had one germline loss-of-function MSH3 or MLH3 mutation and no second hit in the cancer, and were therefore deemed MSH3/MLH3-proficient. One MSH3/MLH3 germline mono-allelic cancer was MSI+ and was excluded. Microsatellite stable (MSS) MSH3/MLH3-WT cancers (n=1,619) had no germline loss-of-function MSH3 or MLH3 mutation, no germline or somatic pathogenic POLEor POLD1 mutation, and no other evidence of mismatch repair deficiency. Microsatellite unstable (MSI+), MSH3/MLH3-WT cancers (n=334) had no germline loss-of-function MSH3 or MLH3 mutation, no germline or somatic pathogenic POLEor POLD1 mutation, and were MSI+ owing to mismatch repair deficiency (MSH2, MLH1, MSH6or PMS2 mutation, or inferred MLH1 hypermethylation). Statistical tests were based on multiple logistic regression, with age, sex, tumour purity and CRC location (proximal or distal colorectum) as co-variables. Further details, including individual indel channel burdens and activities, are in **Supplementary** Table 3.

| Burdens<br>(median, IQR) | MSH3/MLH3-<br>deficient | MSH3/MLH3 germline<br>mono-allelic | MSS MSH3/MLH3-<br>WT | MSI+ MSH3/MLH3-WT | *P* MSH3/MLH3-deficient vs<br>MSH3/MLH3 germline mono-allelic | *P* MSH3/MLH3-deficient vs<br>MSS MSH3/MLH3-WT | *P* MSH3/MLH3-deficient vs<br>MSI+ MSH3/MLH3-WT |
| --- | --- | --- | --- | --- | --- | --- | --- |
| **Total ID** | 4,200 (3,504–5,319) | 1,417 (1,283–1,967) | 1,453 (1,105–1,987) | 142,510 (116,622–168,711) | 0.00300 | 0.00000 | 0.00000 |
| **ID4 Channels** | 1,478 (671–1,827) | 79 (64–105) | 74 (56–95) | 2,549 (1,870–3,409) | 0.01700 | 0.00047 | 0.06700 |
| **1bp Insertion** | 452 (378–685) | 592 (572–769) | 635 (475–874) | 19,008 (14,512–22,812) | 0.98700 | 0.70100 | 0.00000 |
| **1bp Deletion** | 2,127 (1,493–2,527) | 595 (516–887) | 589 (434–822) | 113,302 (92,662–134,562) | 0.00900 | 0.00006 | 0.00000 |
| **≥2bp Insertion** | 102 (61–119) | 73 (67–93) | 72 (52–99) | 2,334 (1,691–3,037) | 0.73600 | 0.53000 | 0.00000 |
| **≥2bp Deletion** | 1,654 (763–2,096) | 145 (101–189) | 131 (100–172) | 7,290 (5,378–9,435) | 0.01400 | 0.00039 | 0.00000 |
| Activities, %<br>(median, IQR) | MSH3/MLH3-<br>deficient | MSH3/MLH3 germline<br>mono-allelic | MSS MSH3/MLH3-<br>WT | MSI+ MSH3/MLH3-WT | *P* MSH3/MLH3-deficient vs<br>MSH3/MLH3 germline mono-allelic | *P* MSH3/MLH3-deficient vs<br>MSS MSH3/MLH3-WT | *P* MSH3/MLH3-deficient vs<br>MSI+ MSH3/MLH3-WT |
| **ID4 Channels** | 34.04 (23.32–40.22) | 5.19 (4.67–5.49) | 4.90 (4.07–6.00) | 1.83 (1.53–2.09) | 0.01000 | 0.00009 | 0.00002 |
| **1bp Insertion** | 10.78 (6.52–27.50) | 42.14 (40.69–46.45) | 44.36 (39.36–48.75) | 13.06 (11.45–15.01) | 0.00100 | 0.00000 | 0.86300 |
| **1bp Deletion** | 46.20 (42.16–52.48) | 41.99 (39.74–45.21) | 40.81 (36.31–46.30) | 80.10 (77.87–81.88) | 0.51000 | 0.01800 | 0.00000 |
| **≥2bp Insertion** | 2.03 (1.24–3.41) | 5.09 (4.54–6.11) | 4.78 (4.09–5.71) | 1.60 (1.32–1.94) | 0.00600 | 0.00024 | 0.58000 |
| **≥2bp Deletion** | 39.03 (26.69–44.44) | 8.44 (7.26–10.91) | 8.80 (7.28–10.83) | 5.22 (4.36–5.85) | 0.01100 | 0.00018 | 0.00002 |

MSH3/MLH3-deficient cancers had a median of ∼4,200 indels, representing a ∼3-fold fold excess over MSS cancers (**Table 2; Supplementary Table 3**), with the burden of ≥2bp deletions at both STRs and sites of microhomology particularly increased (over 12-fold). The burden of 1bp deletions at longer oligoT tracts was also raised in MSH3/MLH3-deficient cancers over MSS tumours, but to a lesser relative extent (∼3.5-fold).

In the 83 individual COSMIC ID channels, activities (relative burden or proportion of all indels) largely mirrored the differences in absolute burden between MSH3/MLH3-deficient and MSS cancers. However, the activities of certain ID channels were higher in MSH3/MLH3-deficient than MSI+ cancers (**Figures 3C & 4; Supplementary Table 3**). This mostly reflected the very large MSI+ 1bp indel burden that lowered the relative burden of other ID channels. Nevertheless, the combination of greater activity and similar burden of longer indels at STRs and sites of microhomology in MSH3/MLH3-deficient cancers suggested a specific role for MSH3 and/or MLH3 in repairing such mutations (**Figure 4; Supplementary Table 3**). We noted that in MSH3/MLH3*-*deficient cancers, CRC tumour suppressor gene spectra included *≥*2bp indels, consistent with the MSH3 or MLH3 deficiency sometimes occurring relatively early in tumorigenesis (**Table 1; Supplementary Table 1**).

**Figure 4.**
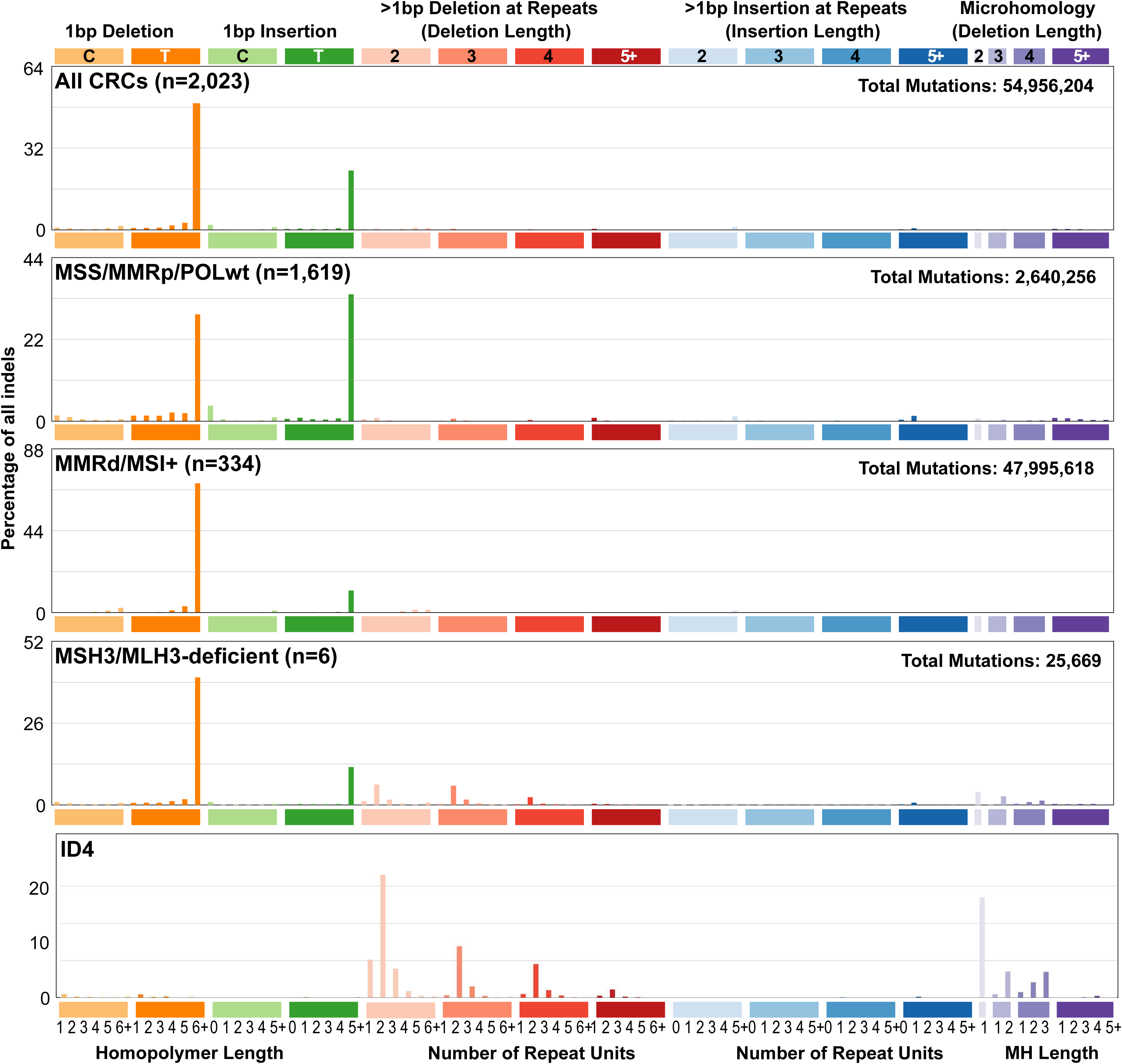
Indel mutation spectra of CRCs and signature ID4. Insertion-deletion mutations from each CRC were assigned to one of the 83 COSMIC ID channels and counts then pooled to produce spectra for all CRCs and for each of the three main groups. Note both the high proportion of T indels, a channel almost absent from ID4, in all CRC types and the signal from an ID4-like ≥2bp indel spectrum specifically in the MSH3/MLH3-deficient tumours ánd in ID4.

Indel mutations in the eight cases with mono-allelic germline *MSH3* or *MLH3* mutations, but no second hits, closely resembled those in the bulk of MSS CRCs (**Supplementary Table 3**). Conversely, and despite the limited samples sizes, the burdens and activity of*≥*2bp deletions were significantly greater in mono-allelic (heterozygous) germline mutation carriers whose CRCs were MSH3/MLH3-deficient (bi-allelic mutant genotype) than the mono-allelic mutant cancers. Overall, the data supported a model in which second hits are required if germline heterozygous *MSH3* or *MLH3* mutations are to have an effect on tumorigenesis.

### MSH3/MLH3 deficiency and COSMIC insertion-deletion signature ID4

Mutational differences between any two cancer types may represent exposures, deficiencies and evolutionary trajectories at different levels, including some factors that are unknown. Previously, extractions of the COSMIC somatic mutational SBS, DBS and ID signatures (https://cancer.sanger.ac.uk/signatures/) had been performed on the entire set of 2,023 CRCs with no knowledge of the patients’ MSH3 or MLH3 status at the time [16]. We replicated the previous ID signature extraction, with identical results.

Whilst the indel mutation spectra of the three main CRC groups (MSS, MSI and MSH3/MLH3-deficient) were dominated by 1bp indels (**Figure 3C**) and hence signatures ID1 and ID2 (data not shown), SigProfileExtractor found that five out of the six MSH3/MLH3-deficient CRCs had the rare COSMIC indel signature ID4, which was absent from all the other 2,017 CRCs (P<0.0000001, Fisher’s exact test). This was the case when either signatures were extracted from the whole set of CRCs, or from the MSH3/MLH3-deficient CRCs alone (**Supplementary Figures 1 & 2**). Since ID4 mostly comprises *≥*2bp deletions at STRs or sites of microhomology (**Figure 4**), this finding was consistent with the mutation spectrum of MSH3/MLH3-deficient tumours. SigProfileExtractor identified de novo three mutational signatures in MSH3/MLH3-deficient CRCs (termed ID83 A, B and C), of which ID83A was decomposed into ID2 and ID4 (combined cosine similarity 0.995), ID83B into ID4 alone (cosine similarity 0.917), and ID83 into ID1 and ID2 (cosine similarity 0.998) (**Supplementary Figure 1**). In the CRCs with ID4 detected, its activity (contribution to the burden of all indels) ranged from 0.227 to 0.477. The simple spectrum of ≥2bp indels only (COSMIC ID channels 25-83) in the combined mutations from the six MSH3/MLH3-deficient CRCs had strong cosine similarity (0.913) to ID4 (**Figure 4**). Bar the single ID4-negative case, the decomposed ID mutation spectra of individual MSH3/MLH3-deficient CRCs, derived by fitting a combination of de novo extracted signatures to COSMIC reference signatures, had close visual resemblance to ID4 outside the 1bp indel channels, with cosine similarity above 0.9 in each case (**Supplementary Figure 2**).

Since our results suggested that MSH3/MLH3-deficient CRCs had similar features to the bulk of MSS cancers, apart from more deletions in the former, we also determined the mutation spectrum in the pooled MSH3/MLH3-deficient CRCs having subtracted mutations present in the bulk of MSS cancers (**Supplementary Figure 3**). This again indicated strong similarity of the *≥*2bp indel channels to ID4, but there was also an increase in 1bp deletions at oligoT tracts (essentially signature ID2). This excess was smaller than that for *≥*2bp indels (about 4-fold versus 12-fold), and was especially apparent for tracts comprising five or more Ts (**Supplementary Table 3**). It was not revealed by signature extraction because ID2 is almost ubiquitous in CRCs. Nevertheless, the data suggested that MSH3 and MLH3 have a role in repairing these shorter deletions [20, 21, 22], perhaps because the indel loops caused by replication slippage distort the double helix in a similar way to longer deletions and are thus identifiable by MSH2-MSH3 heterodimers.

Why one MSH3/MLH3-deficient CRC was ID4-negative was unclear. Unlike its five counterparts, the MSH3/MLH3-deficient CRC was from the most proximal part of the colorectum (caecum), which is subject to quantitatively different mutational processes from the rest of the colorectum, probably including more 1bp indels (Cornish et al, 2024). We speculate that this influenced the resulting somatic mutation spectrum, although we accept that other explanations for the lack of ID4 in this cancer are possible (for example, the second hit at MLH3 might have occurred at a relatively late stage of cancer evolution, and genotoxic therapy might have been used but not recorded accurately). Like the other MSH3/MLH3-deficient cancers, the ID4-negative tumour had some ID4-like features compared with the set of MSS cancers, including an excess of ≥2bp deletions (burden=284 v median burden MSS=130; P=0.041, one-tailed rank test) and ID4 channel mutations (burden=147 v median burden MSS =74; P=0.043, one-tailed rank test).

### Germline MSH3, and probably MLH3, loss-of-function variants predispose to colorectal cancer

There are occasional reports in the literature of heterozygous germline *MSH3* or *MLH3* mutations in CRC cases [12, 23, 24, 25, 26, 27, 28] (**Supplementary Table 4**). The largest such study was that of Salo-Mullen *et al* [29], who found 13 germline *MSH3* mutation carriers (including one with bi-allelic variants) in a pan-cancer data set. However, there was not an increased frequency of these variants in cancer cases over controls. Another study found suggestive evidence that an uncommon missense *MSH3* variant, rs200819607, predicted to be functionally non-pathogenic [30], was over-represented in CRC cases. However, we found no evidence of a similar association in our data (1/3,025 rs200819607 heterozygote case *versus* 3/14,409 controls in 100kGP, and 1/6,057 cases *versus* 72/147,694 controls in UKB (OR=0.66, Passoc(meta)= 0.584. The various previous studies have not unreasonably prompted suggestions that *MSH3* and *MLH3* heterozygotes could be at increased CRC risk. However, previous studies have mostly lacked somatic mutation data and have often comprised only one or two *MSH3* and *MLH3* heterozygotes. Consequently, it has not previously been possible to show that heterozygotes are predisposed to CRC or other cancer types.

Since there were germline heterozygotes in our data sets who did not develop CRC or whose CRCs did not have second hits, it was likely that penetrance of germline *MSH3* and *MLH3* mutations was incomplete. We reasoned that it was necessary to determine formally whether heterozygous germline LoF *MSH3* or *MLH3* mutations predispose to CRC. In an analysis of over 11,000 colorectal tumour cases and more than 450,000 cancer-free controls from the 100kGP, CORGI, UK Biobank and AllofUs studies (Figure 5; Methods), we found that *MSH3* heterozygotes had a significantly increased risk of CRC and/or polyps (OR=2.155, *P*=6.6x10^-5^; Figure 5). There was also an association between germline *MLH3* heterozygosity and CRC risk (OR=1.553, *P=*0.028).

**Figure 5.**
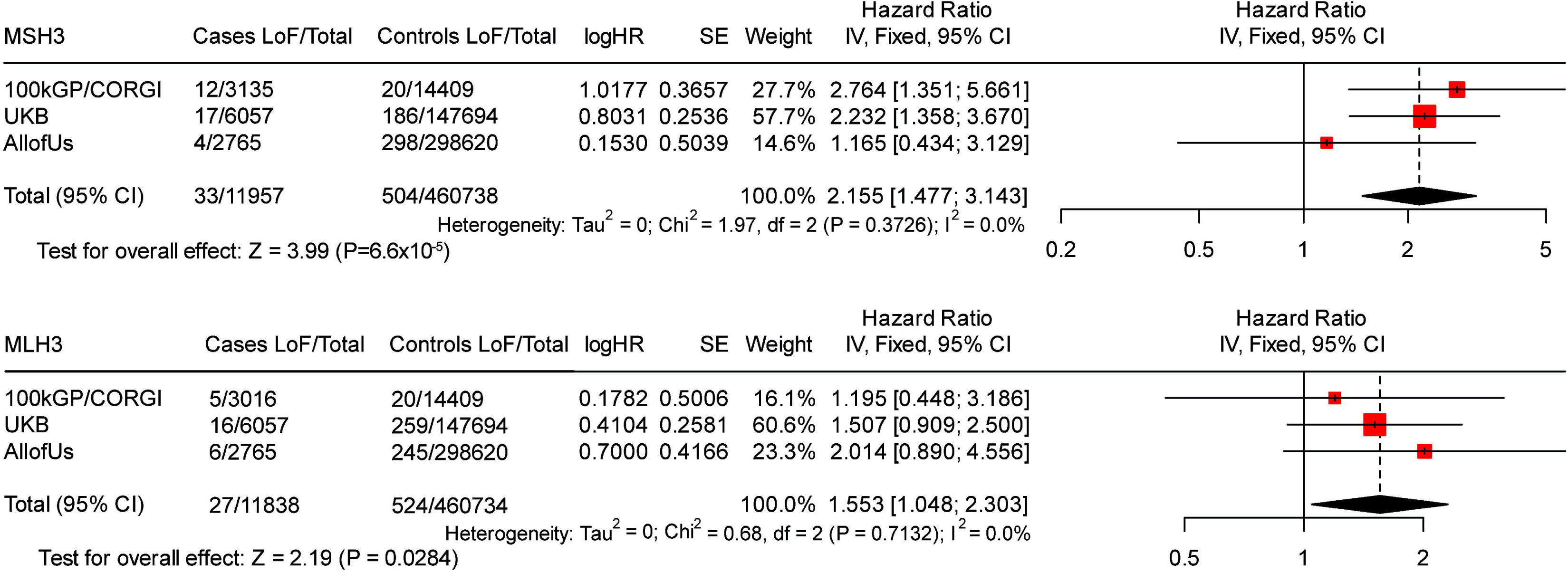
Meta-analyses of case-control association studies, comparing the frequency of mono-allelic, germline LoF *MSH3* and *MLH3* variants in colorectal tumour cases and controls from 100kGP+CORGI, UKB and AllofUs. Association statistics with corresponding 95% confidence intervals (CIs) and inter-study heterogeneity metrics (I² and P_het_) are shown.

### Germline MSH3 and MLH3 variants and extra-colonic cancers

We broadened our search for pathogenic germline *MSH3* and *MLH3* variants to cancers other than CRC using two sets of 100kGP patients: (i) 13,588 with genome sequencing of paired tumour and normal tissue from the Pan-Cancer Domain (excluding CRC) and (ii) 1,928 individuals from the Rare Disease domain, with tumour syndromes other than familial colorectal cancer and multiple bowel polyps. In the 15,516 patients with extra-colonic malignancies, no bi-allelic mutation carriers were found, and 47 patients carried germline heterozygous LoF mutations (20 *MSH3*, 27 *MLH3*) (**Supplementary Tables 2 & 5**). There was neither an overall increased risk of extra-colonic tumours in general in heterozygotes (OR=0.977, *P=*1.00, Fisher’s exact), nor of any specific type of tumour (details not shown). We also searched for the presence of ID4 in relation to *MSH3* and *MLH3* genotype in the 10 most common types of extra-colonic carcinoma in 100kGP (**Supplementary Tables 5 & 6**). Whilst only two breast carcinoma cases were *MLH3* heterozygotes, both cancers had second hits by LOH, had relatively high levels of 1bp and ≥2bp deletions, and were positive for ID4, which was present in less than 1% of all breast cancers overall (*P=*1.79x10^-4^, Fisher’s exact; **Supplementary Tables 5 & 6; Supplementary Figure 2**).

## Discussion

In this study, we initially confirm published findings that bi-allelic germline *MSH3* mutations cause multiple polyps and CRC, and provide evidence in support of the limited existing data that bi-allelic *MLH3* mutations have a similar effect [6, 10]. We then show for the first time that mono-allelic (heterozygous) germline *MSH3* mutations, and probably *MLH3* mutations, predispose, to CRC, conferring about 1.5-2.2-fold increased risk (∼14% estimated lifetime risk). Somatic second hits can lead to *MSH3* or *MLH3* inactivation and thus to relatively mild hypermutation, most strikingly taking the form of increased ≥2bp deletions, but also including higher burdens of 1bp deletions. This is likely to cause an increased chance of colorectal carcinogenesis. MSI is typically absent from MSH3/MLH3-deficient cancers, which are thus formally MMRd but MSS.

Where somatic loss of *MSH3* or *MLH3* does occur in mono-allelic mutation carriers owing to second hits, these MSH3/MLH3-deficient cells may persist and the hypermutation may promote tumorigenesis. However, it is also clear that ‘sporadic’ CRCs can arise in germline mutation carriers, without *MSH3* or *MLH3* dysfunction playing any role. We postulate that in these cases, ‘second hits’ may not have occurred, or cells with a second hit did not persist, because they only provided a very small and indirect selective advantage. As a result, disease penetrance is incomplete in mono-allelic *MSH3* or *MLH3* carriers.

There is clear evidence that mutational signatures act as phenotypes that mediate increased CRC risk in DNA repair deficiencies such as Lynch syndrome (multiple ID and SBS signatures), polymerase proofreading polyposis (SBS10), *MUTYH*polyposis (SBS36) and *MBD4-*associated neoplasia (SBS1). ID4 may play an equivalent role in MSH3/MLH3-deficient CRCs. Whilst we found no statistical association between *MSH3* or *MLH3* germline heterozygosity and the risk of extra-colonic cancers, we did find two breast cancers from carriers of heterozygous germline *MLH3* mutations. These tumours had acquired somatic second hits and showed both increased indels and ID4. A small raised risk of breast cancer might thus accompany the increased risk of CRC in these patients.

ID4 includes almost no 1bp indel channel mutations, and thus ID4 appears not to be a simple reflection of MSH3/MLH3 deficiency. It does remain possible that the increased 1bp deletion burden in MSH3/MLH3-deficient cancers reflects another process, such as increased replication rates, but there is prior *in vitro* evidence that MSH3 and MLH3 participate in the repair of 1bp deletions [20, 21, 22]. Instead, ID4 may simply reflect, with high accuracy, the ≥2bp indels caused by deficient MSH3 or MLH3, whereas the 1bp indels are mostly captured by the reference signatures ID2 and ID1, which comprise over 80% of all indels in tumours lacking canonical MMR (mostly MLH1-deficient). We also note that ID4 is present in a small proportion (typically <1%) of other 100kGP cancers without evidence of MSH3 or MLH3 deficiency. Whilst we could not determine whether those tumours had other causes of MSH3 or MLH3 loss, such as epimutations, those observations leave open the possibility that ID4 can have multiple origins.

Consistent with multiple origins for ID4, Reijns *et al* [31] have reported its presence secondary to combined loss of Rnaseh2b and p53 function in cell and mouse models, in a study to examine the consequences and removal of ribonucleotides incorporated into DNA. In human chronic lymphocytic leukaemias (CLLs) with bi-allelic *RNASEH2B*deletions, Reijns *et al* found a raised burden of 2-5bp deletions, but they did not find that ID4 itself was associated with *RNASEH2B*deficiency. Reijns *et al* posited that the 2-5bp deletions were caused by topoisomerase 1 (TOP1) preferentially cleaving DNA at ribonucleotides that could not be removed from DNA in RNASEH2B-deficient cells. Cleavage was thought to target the motif TN_n_T especially (where N represents any nucleotide and n represents any repeated number of that nucleotide). However, whilst the ≥2bp indel spectra of the Reijns *et al* mice and our MSH3/MLH3-deficient human cancers both resemble ID4 (cosine similarity 0.913 and 0.866 respectively), their resemblance to each other is more limited (cosine similarity 0.668; **Supplementary Figure 4**). Whilst some of this difference may be due to inter-specific variation, it does suggest that MSH3/MLH3 deficiency and RNASEH2B deficiency have fundamental differences. Further investigation will be required to explore the possible relationships between ID4, MSH3/MLH3 deficiency and the ID-TOP1 signature (TN_n_T>T), but we believe it possible that a major role of MSH3 and MLH3 could be to repair TOP1-mediated deletions, especially if they occur as part of DNA replication. RNASEH2B and MSH3/MLH3 deficiency might thus converge on a similar mutational spectrum that is ID4. To support this possibility, we annotated the deletion mutations in our MSH3/MLH3-deficient and MSS bulk CRCs into those at or outside motifs prone to TOP1 cleavage, using the script employed by Reijns *et al* (https://git.ecdf.ed.ac.uk/ID-Top1/wgs_variant_analyses/-/blob/master/utils/utils.py?ref_type=heads). Considering all deletions, about 85% occurred at TnT compliant sites in MSS CRCs, whereas the equivalent for MSH3/MLH3-deficient CRCs was 93% (P<10-5, Fisher’s exact test, odds ratio 2.52). For ≥2bp indels specifically, the equivalent metrics were 77% and 94% (P<10-5, Fisher’s exact test, odds ratio 4.34). Thus, mutations arising from MSH3/MLH3 deficiency seemed to be enriched for longer indels at TnT compliant sites, consistent with a particular role for MSH3 and MLH3 in repairing TOP1-mediated damage.

Our analysis has limitations, some of which are intrinsic to the study of rare genetic variants that have an intermediate magnitude of effect on disease risk. These issues affect many studies of the notorious ‘missing heritability’ of human disease [32]. Our CRC cases, for example, have some bias towards familial, early-onset or multi-tumour phenotypes. These enrichments probably empower the detection of true associations, but could lead to risk quantitation that is inaccurate for the general population. Furthermore, whilst the availability of copious comparison data, such as the many cancer genomes from 100kGP, can lead to impressive and plausible association statistics, such as those between MLH3 deficiency and ID4 in breast cancer, we must bear in mind that only two such tumours had both of these molecular features in our study. Nevertheless, even with incomplete penetrance, we note that the predicted effect of germline *MSH3* heterozygosity on population CRC burden is much greater than that of bi-allelic *MSH3* mutations. A further issue is that archival cancer specimens are not available within 100kGP to allow ancillary analyses, such as immunohistochemistry, to be performed.

Overall, we have validated the hypothesis that germline *MSH3* mutations predispose to colorectal tumours not only in the bi-allelic state, but also as heterozygotes. Germline *MLH3* mutations probably have similar effects. The relative risk in heterozygotes is about two and thus *MSH3* and *MLH3* could be said to be moderate risk CRC genes, among a small set of genes of this type with variable supporting evidence [33]. The CRCs resulting from germline *MSH3* or *MLH3* mutations resemble CRC-specific forms of both Lynch syndrome and cMMRd in that those tumours are MMR-deficient and have a specific hypermutator phenotype, comprising excess ≥2bp deletions resembling ID4 and a smaller relative excess, but similar extra burden, of 1bp deletions resembling ID2. However, those tumours are classed as microsatellite-stable, [19] principally because their burden of 1bp indels is 5-fold less than those of MSI+ tumours with *MLH1* inactivation, and their total TMB still resembles that of MSS cancers. In these respects, the effects of germline *MSH3* and *MLH3* mutations are lower than those of *MSH3* and *MLH1,* as is also apparent to a lesser extent for *PMS2* [5, 34]. Given that Lynch syndrome is ideally a molecular diagnosis and that genotype-phenotype differences among the four genes are well described [35], it is at least arguable that *MSH3* and *MLH3* should be added to *MSH2, MLH1, MSH6*and *PMS2* as causes of both Lynch syndrome and cMMRd.

## Methods

### Clinicaldata

For 100kGP, ethical approval was provided by the HRA Committee East of England – Cambridge South research ethics committee (REC Ref 14/EE/1112). 100kGP patient clinical data for CRC cases were obtained through Participant Explorer within the Genomics England research environment, which is linked to NHS patient data *via* hospital inpatient records, national cancer registries, prescribing of chemotherapy and radiotherapy, UK Office of National Statistics, death statistics and national histology records. In addition to CRC and/or multiple polyp cases, 100kGP recruited individuals with other cancer types, rare phenotypes of an unexplained, but potentially genetic origin, and unaffected family members.

Patient medical history, family history and tumour data were collected directly from CORGI participants, who provided full informed consent (UK Research Ethics Committee approvals 06/Q1702/99 and 17/SC/0079). Patient recruitment in CORGI was enriched for cases with early-onset CRC, a family history of CRC or multiple polyps.

We applied genetic ancestry and kinship filters to select for Europeans and unrelated individuals using the same threshold values as the other cohorts (>0.99 and <0.01625 respectively), leaving 3,025 100kGP and CORGI cases. We pre-identified as “controls” 14,409 unrelated 100kGP participants with no personal or close family history of cancer, or of a condition potentially associated with cancer [14].

For UKB CRC cases, data were obtained under UK Biobank project ID 155012. All individuals with a current diagnosis or personal history of colorectal cancer or polyps were identified based on International Classification of Diseases ICD-10 codes C180, C182-189, C19 or C20 using data field 40006 or ICD-9 codes 1530-1534 or 1536-1541 using data field 40013. We further included individuals with tumour behaviour described as “malignant, primary site” in data field 40012 and with histology information available in data field 40011 to confirm the CRC diagnosis. Tumours that were self-reported only (provided in data field 20001) could not be validated and were therefore excluded.

For UKB controls, we excluded any individuals with an ICD-9 or ICD-10 cancer, or self-reported cancer diagnosis (provided in data fields 40013, 40006 and 20001 respectively), or reports of colorectal polyp(s) based on ICD-10 codes D12, D120, D121, D122, D123, D124, D125, D126, D127, D128 or D129) in data fields 41202 and 41204, or ICD9 codes 211, 2111, 2112, 2113, 2114, 2115, 2116, 2117, 2118 or 2119 in data fields 41203 and 41205, or code 1460 in data field 20002. We also removed all individuals with reported history of cancer in either parent or any sibling of the cancer types with data available (breast, lung, bowel or prostate cancers). We then applied genetic ancestry and kinship filters as above. The ages of UKB controls were similar to those of cases (medians 56 versus 63 respectively).

Carriers of germline *MSH3* and *MLH3* mutations of self-reported White ancestry were also assessed from short read germline WGS data on a non-patient-identifiable basis as part of registered project 68572 in the All of Us study. The data set for association analysis comprised 2,765 CRC cases and 298,620 cancer-free controls.

### Association analysis of CRC risk

Frequencies of putative pathogenic germline *MSH3* and/or *MLH3* mutations in cases (CRC or multiple polyps) and controls (cancer-free) were compared using logistic regression analysis. Odds ratios and association statistics were reported. Meta-analysis was performed using STATA v19.5. Association analysis results are presented here without sex and age as co-variables, for the following reasons. The cases in CORGI and the 100kGP rare disease and familial CRC domains were enriched for familial or early-onset CRC, and/or for the presence of multiple adenomatous polyps. This strategy generally increases the statistical power to detect associations with rare variants [36]. Whilst inclusion of sex and age as co-variables in genetic association analysis can generally improve study power, the use of co-variables in the analysis of phenotypically enriched cases can reduce power and introduce spurious associations that obscure the genetic associations of interest [37]. This was specifically manifest here in strong associations between case status and both younger age (enrichment for genetic aetiology) and female sex (probably owing to higher chances of recruitment) in CORGI. These contrast with the established population associations between CRC and both older age and male sex. Of note, the associations between CRC and younger age and female sex were not present in UKB, which is a volunteer-based population cohort study with a restricted age range and no particular selection for cancer or other phenotypes.

### Pan-cancer (CRC-excluded) analysis

Sets of patients with malignancies other than CRC were obtained from 100kGP using equivalent criteria to those used for CRC cases, partly to evaluate any effect of germline heterozygosity for *MSH3* or *MLH3* on Lynch syndrome cancers other than CRC, including the presence of ID4 in those cancers. We also wished to make a comparison with the results of Vali-Pour *et al* [38] who had assigned an unknown set of principally missense changes of uncertain pathogenicity in *MSH3* and *MLH3* (combined frequency ∼1%) to somatic indels of ≥2 bases. However, we could not identify any relevant germline variants at about 1% combined frequency, and our loss-of-function variants’ allele frequencies were an order of magnitude less (∼0.1%), hence we were unable to make the comparison. We proceed to explore the association between *MSH3* or *MLH3* germline heterozygotes and pan-cancer (CRC-excluded) risk in 100kGP.

### Whole-genome sequencing of constitutional and tumour DNA from patients with CRC and/or multiple colorectal polyps

Whole genome sequencing of constitutional genomic DNA (gDNA) in 100kGP was performed using Illumina 75bp paired end sequencing. Details of the data quality control and analysis pipelines are as reported [16]. In brief, WGS was performed on DNA from peripheral blood from all participants to a median depth of ∼30X. CORGI patient WGS data were obtained and processed similarly, and combined with the 100kGP data for analysis as a single data set.

Frozen tumours (100kGP CRCs) were sequenced to a median depth of ∼100X. Reads were mapped to GRCh38 with the ISAAC aligner and germline variants called with Starling. Small somatic variants were called with Strelka and structural variants with MANTA. msiNGS [19] was used to call microsatellite instability, supplemented in a small number of equivocal cases by mutation burden assessment and COSMIC signature analysis. Somatic copy number and loss-of-heterozygosity (LOH) analyses were performed using the Battenberg program, supplemented by the Sequenza program where Battenberg produced equivocal results.

FASTQ files were mapped to build 38 (hg38) of the human genome reference using an in-house pipeline making use of SAMtools, BEDtools, fastqutils, BWA aln, STAMPY and the picard suite of tools (http://broadinstitute.github.io/picard/). Deduplicated BAM files were then submitted to Genome Analysis Tool Kit (GATK) pre-processing for variant discovery steps: IndelRealigner, BaseRecalibrator, PrintReads. HaplotypeCaller was used in GVCF mode to call variants in per sample mode using the analysis ready BAMs. CombineGVCFs was used to generate per chromosome multi-sample GVCFs. Variants were then joint called in discovery mode using a minimum phred-scaled Q score confidence threshold of 10 for emission and of 30 for high-confidence calling. Variant Quality Score Recalibration (VQSR) was used to filter the raw variant calls with all variants in the 99.9% and above tranches being retained. Variant tools (VT)(50) was used in smart mode to decompose variants to bi-allelic sites and variant annotation was performed using the Ensembl Variant Effect Predictor (VEP) framework. Specific analyses of germline and somatic mutations, copy number changes, LOH and measures of mutation burden were essentially as reported by Cornish *et al* [16].

### Mutational signatures

Mutational signatures had previously been computed and reported as part of the 100kGP CRC landscape study [16]. We replicated this work as part of this study. In addition to the default Strelka filters, Cornish *et al* [16] had applied the following exclusion filters for defining a set of high confidence somatic variants:

- Variants with a germline allele frequency (AF) >1% in the full Genomics England dataset
- Variants with a population germline AF >1% in the gnomAD database [39]
- Somatic variants with frequency >5% in the Genomics England cancer dataset. A 5% cut-off was chosen based on the frequency of recurrent non-synonymous variants in Cancer Gene Census genes [40]
- Variants overlapping simple repeats as defined by Tandem Repeats Finder [40]
- Indels in regions with high levels of sequencing noise where >10% of the base calls in a window extending 50bps either side of the indel have been filtered out by Strelka due to the poor quality
- Indels within 10bp of 100kGP or Gnomad v3 germline indel with AF >1%
- Variants in regions of poor mappability where the majority of overlapping 150bp reads do not map uniquely to the variant position
- SNVs resulting from systematic mapping and calling artefacts present in both tumour and normal 100kGP sample sets

For the last category (systematic mapping and calling artefacts), we tested whether the ratio of tumour allele depths at each somatic SNV site was significantly different to the ratio of allele depths at this site in a panel of normal samples (PoN) using Fisher’s exact test. The PoN was composed of a cohort of 7,000 non-tumour genomes from the Genomics England dataset. At each genomic site only individuals not carrying the relevant alternative allele were included in the count of allele depths. The mpileup function in bcftools (v1.9) was used to count allele depths in the PoN. To replicate Strelka filters duplicate reads were removed and quality thresholds set at mapping quality ≥5 and base quality ≥5. All somatic SNVs with Fisher’s exact test phred score <80 were filtered, with the threshold determined by optimising precision and recall calculated from a TRACERx truth set [41].

Single-base-substitution (SBS), doublet-base-substitution (DBS) and insertion and deletion (indel; ID) signatures were extracted *de novo* and related to known COSMIC signatures (v3.2) using SigProfilerExtractor 75 [16]. SBS, DBS and ID signatures were extracted using random initialization, 500 NMF replicates, and between 10,000 and 1,000,000 NMF iterations. We assumed the presence of 1-30 SBS signatures (minimum_signatures and maximum_signatures parameters respectively), 1-15 DBS signatures, and 1-10 ID signatures during analysis, with the final number of *de novo* signatures determined by an optimum balance of information, parsimony and statistical support [42]. Default settings were used for all other parameters. For each tumour, we calculated burdens of mutations of any one type (e.g. assigned to a COSMIC channel based on the specific changes or to a signature by SigProfilerExtractor), activities (burdens relative to all mutations of a specific type) and presence/absence (identified as present by SigProfilerExtractor when fitting COSMIC reference signature to *de novo* extracted signatures). We note that signature extraction is a very innovative, powerful and useful method, but potential issues are inherent and must be borne in mind: for example, signatures are assessed based on activities and thus the presence of one mutational process can reduce the activity of a concomitant process, hence obscuring the latter.

### UK Biobank and All of Us studies: participant WGS of constitutional DNA

DNA was extracted from peripheral blood. Molecular and clinicoathological data were extracted from the UK Biobank RAP (https://www.ukbiobank.ac.uk/media/dovbae03/uk-biobank-final-whole-genome-sequencing-release-faqs_v1-0.pdf) or from the AllofUs research hub (https://www.researchallofus.org/).

### Swedish CRC cases

Germline *MSH3* and *MLH3* variants and somatic mutations were derived from the set of CRCs with WGS reported by Nunes *et al* [43]. These cases are listed in **Supplementary Table 2**, but the data set was too small to contribute to the association analysis. Moreover, none of these cancers had second hits and thus they did not contribute to the assessment of hypermutation or ID4.

### Germline variant annotation and pathogenicity assignment

We used ClinVar pathogenicity annotations for germline and somatic variants of interest, where available. We also inspected sites of mutations in the Integrated genomics viewer (IGV), in order to exclude locations with recurrently poor quality sequence in either gene. In practice, since almost all missense *MSH3* and *MLH3* mutations were classed as benign, probably benign or of uncertain, doubtful or conflicting pathogenicity, we excluded all missense changes, unless they potentially affected mRNA splicing. We thus initially regarded only loss-of-function (LoF, *i.e.* frameshift or stop-gained) variants as pathogenic. A search for missense germline mutations targeting critical amino acids in functionally important domains as identified by Uniprot and BLAST revealed <5 such changes in our data set, and we did not include them in our analysis. SpliceAI was used to assess the pathogenicity of splice region substitution and indel variants. Recommended cut-offs (probability of change in splice status ≥ 0.8) were used to score a variant as probably pathogenic. gnoMAD v3.1.2 and v4.1.0/non-UKB databases were utilised to provide informal comparisons with our data, given the limitations that colorectal and other tumour statuses were not available, compound heterozygotes could not be identified, and participant ancestry was unknown.

### RNA sequencing

RNAseq was performed on total RNA extracted from an additional sample taken from 265 CRCs that had undergone WGS. Sequencing methods, outcome metrics, data analysis methods inc. gene expression levels, etc. For the patient with unexplained ID4, PCA was performed to confirm that the tumour was not an outlier for any reason and sufficient tumour purity was confirmed by confirming ready detection of somatic driver mutations previously found by WGS (*APC, IDH2, FBXW7*) in RNAseq BAM files visualised in the Integrated Genomics Viewer. In detail, RNA sequencing was performed using TruSeq Stranded Total RNA Library Prep kit (Illumina) with H/M/R Gold probes. Pooled libraries were sequenced using 75bp paired end reads on HiSeq4000 aiming for 50 million reads. Fastq reads were quality checked by trimming for low-quality ends and clipping adapter sequences if present, using Trimmomatic (v.0.36). QC processed reads were then mapped to the human genome (GRCh38) and transcriptome (Ensembl release 87) using STAR aligner (v. 2.6.2b) two-pass mode. Gene level count data thus obtained were subsequently normalised using the ’voom’ method and differential gene-expression comparisons using the empirical Bayes method implemented in limma package (v.3.44.1) in R (v.3.4.0).

### Availability of data and materialss

Data from the National Genomic Research Library (NGRL) used in this research are available within the secure Genomics England Research Environment. Access to NGRL data is restricted to adhere to consent requirements and protect participant privacy. Data used in this research include cases from the cancer domains, rare diseases (familial colorectal cancer and multiple polyps, and other domains (controls). Data from these participants is available through LabKey in the NGRL, subject to access being provided to approved researchers who are members of the Genomics England Research Network, to institutional access agreements, and to research project approval under participant-led governance. For more information on data access, visit: https://www.genomicsengland.co.uk/research.

This research was also conducted using the UK Biobank Resource under application numbers 19655 and 155012. UK Biobank data are available through application to the UK Biobank (https://www.ukbiobank.ac.uk/use-our-data/apply-for-access/), with permission of UKB’s Research Ethics Committee.

Other data (e.g. from CORGI study. Swedish study) will be made available by the lead researchers to collaborating researchers, subject to ethical permissions and formal agreement.

All of Us data are available to registered researchers through application https://www.researchallofus.org/.

## Other information

### Author contributions

Planned study and wrote manuscript – IS, IT, JFT; analysed data – IS, KS, JW, JFT, AA, ST, IT, GG, JW, CP, AG Provided data – AI, BG, TS, CP. Guarantor – IT

## Data Availability

Data from the National Genomic Research Library (NGRL) used in this research are available within the secure Genomics England Research Environment. Access to NGRL data is restricted to adhere to consent requirements and protect participant privacy. Data used in this research include cases from the cancer domains, rare diseases (familial colorectal cancer and multiple polyps, and other domains (controls). Data from these participants is available through LabKey in the NGRL, subject to access being provided to approved researchers who are members of the Genomics England Research Network, to institutional access agreements, and to research project approval under participant-led governance. For more information on data access, visit: https://www.genomicsengland.co.uk/research.This research was also conducted using the UK Biobank Resource under application numbers 19655 and 155012. UK Biobank data are available through application to the UK Biobank (https://www.ukbiobank.ac.uk/use-our-data/apply-for-access/), with permission of UKB's Research Ethics Committee. Other data (e.g. from CORGI study. Swedish study) will be made available by the lead researchers to collaborating researchers, subject to ethical permissions and formal agreement. All of Us data are available to registered researchers through application https://www.researchallofus.org/.

## Acknowledgements

This work made use of the resources provided by the Edinburgh Compute and Data Facility (ECDF) (http://www.ecdf.ed.ac.uk/). We gratefully acknowledge the participants of the National Genomic Research Library (NGRL), whose contributions made this research possible (National Genomic Research Library, Genomics England (2024). https://doi.org/10.6084/m9.figshare.4530893). Secure access to the NGRL under project ID 26 was provided by Genomics England, which delivers the NGRL in partnership with NHS England, and is wholly owned by the UK Department of Health and Social Care. The NGRL contains participants’ health data collected by the NHS as part of their care, along with samples and data from their participation in research, for which fully informed consent has been obtained. This includes genomic and clinical data provided through the NHS Genomic Medicine Service, as well as data obtained through research studies, including the 100,000 Genomes Project and the Generation Study, both of which are delivered in partnership with the NHS, and from other research cohorts involving external collaborators. This research has been also been conducted using the UK Biobank Resource under application numbers 19655 and 155012. We also thank the National Institutes of Health’s *<u>AllofUs</u>*<u>Research Program</u> for making available the participant data [and/or samples and/or cohort] examined in this study. Finally, we acknowledge the invaluable help of all participants in all studies (CORGI, 100kGP, UKB and AllofUs), especially their provision of data and samples.

## Ethics approval and consent to participate

Ethical approval for collection and analysis of 100kGP patient samples was obtained from the HRA Committee East of England–Cambridge South Research Ethics Committee (REC reference 14/EE/1112). UK Biobank was approved by North West - Haydock Research Ethics Committee 21/NW/0157, IRAS project ID: 299116. The CORGI and CORGI 2 studies were approved by Southampton and SW Hampshire and South Central Research Ethics Committees in the UK with references 06/Q1702/99 and 17/SC/0079. Ethical oversight was provided by the AllofUs IRB (https://allofus.nih.gov/about/who-we-are/institutional-review-board-irb-of-all-of-us).

## Patient and Public Involvement

Members of the public are centrally involved in the design and implementation of large studies including 100kGP, UKB and AllofUs. Members of the public are part of the research ethics committee who reviewed all the documentation. Including the protocol, for the CORGI study. We provide all participants in CORGI with email addresses for the study team so they can report any feedback. Outside the REC review and individua; feedback, patients/public were not involved in the design of the CORGI study, but we consult CORGI participants individually regarding how they value knowledge of a genetic variant within their family and its ability to guide appropriate screening.

## References

1 Boland CR, Lynch HT. The history of Lynch syndrome. Fam Cancer 2013;12:145–57.

2 Ricciardone MD, Ozçelik T, Cevher B, Ozdağ H, Tuncer M, Gürgey A, et al. Human MLH1 deficiency predisposes to hematological malignancy and neurofibromatosis type 1. Cancer Res 1999;59:290–3.

3 Lipkin SM, Wang V, Jacoby R, Banerjee-Basu S, Baxevanis AD, Lynch HT, et al. MLH3: a DNA mismatch repair gene associated with mammalian microsatellite instability. Nat Genet 2000;24:27–35.

4 Brisbin A, Weissman MM, Fyer AJ, Hamilton SP, Knowles JA, Bustamante CD, et al. Bayesian linkage analysis of categorical traits for arbitrary pedigree designs. PLoS One 2010;5:e12307.

5 Will O, Carvajal-Carmona LG, Gorman P, Howarth KM, Jones AM, Polanco-Echeverry GM, et al. Homozygous PMS2 deletion causes a severe colorectal cancer and multiple adenoma phenotype without extraintestinal cancer. Gastroenterology 2007;132:527–30.

6 Adam R, Spier I, Zhao B, Kloth M, Marquez J, Hinrichsen I, et al. Exome Sequencing Identifies Biallelic MSH3 Germline Mutations as a Recessive Subtype of Colorectal Adenomatous Polyposis. Am J Hum Genet 2016;99:337–51.

7 Aelvoet AS, Hoekman DR, Redeker BJW, Weegenaar J, Dekker E, van Noesel CJM, et al. A large family with MSH3-related polyposis. Fam Cancer 2023;22:49–54.

8 Villy MC, Masliah-Planchon J, Schnitzler A, Delhomelle H, Buecher B, Filser M, et al. MSH3: a confirmed predisposing gene for adenomatous polyposis. J Med Genet 2023;60:1198–205.

9 Koi M, Leach BH, McGee S, Tseng-Rogenski SS, Burke CA, Carethers JM. Compound heterozygous MSH3 germline variants and associated tumor somatic DNA mismatch repair dysfunction. NPJ Precis Oncol 2024;8:12.

10 Olkinuora A, Nieminen TT, Mårtensson E, Rohlin A, Ristimäki A, Koskenvuo L, et al. Biallelic germline nonsense variant of MLH3 underlies polyposis predisposition. Genet Med 2019;21:1868–73.

11 Johannesen KM, Karstensen JG, Rasmussen A, Scott EAH, Birkedal U, Hansen TVO, et al. A Novel Case of Biallelic MLH3 Variants in a Patient With Rectal Cancer and Polyps. Clin Genet 2025;107:480–2.

12 Yang M, Zhu L, Xu D, Weng S, Fang X, Dong C, et al. How should we correctly interpret biallelic germline truncating variant of MLH3 in hereditary colorectal cancer? Genet Med 2019;21:2650–1.

13 Palles C, West HD, Chew E, Galavotti S, Flensburg C, Grolleman JE, et al. Germline MBD4 deficiency causes a multi-tumor predisposition syndrome. Am J Hum Genet 2022; 109:953–60.

14 Palles C, Freeman-Mills L, Arbe-Barnes E, Feeley N, Chegwidden L, Curley H, et al. Comparison between germline and somatic loss-of-function RNF43 mutations reveals different genotype-phenotype associations and provides insights into the genetic mechanisms of colorectal tumourigenesis. Gut 2025.

15 Howarth K, Ranta S, Winter E, Teixeira A, Schaschl H, Harvey JJ, et al. A mitotic recombination map proximal to the APC locus on chromosome 5q and assessment of influences on colorectal cancer risk. BMC Med Genet 2009; 10: 54.

16 Cornish AJ, Gruber AJ, Kinnersley B, Chubb D, Frangou A, Caravagna G, et al. The genomic landscape of 2,023 colorectal cancers. Nature 2024; 633:127–36.

17 Young J, Leggett B, Ward M, Thomas L, Buttenshaw R, Searle J, et al. Frequent loss of heterozygosity on chromosome 14 occurs in advanced colorectal carcinomas. Oncogene 1993;8:671–5.

18 Cross W, Kovac M, Mustonen V, Temko D, Davis H, Baker A-M, et al. The evolutionary landscape of colorectal tumorigenesis. Nature Ecology and Evolution 2018; In press.

19 Salipante SJ, Scroggins SM, Hampel HL, Turner EH, Pritchard CC. Microsatellite instability detection by next generation sequencing. Clin Chem 2014;60:1192–9.

20 Chen PC, Dudley S, Hagen W, Dizon D, Paxton L, Reichow D, et al. Contributions by MutL homologues Mlh3 and Pms2 to DNA mismatch repair and tumor suppression in the mouse. Cancer Res 2005;65:8662–70.

21 Flores-Rozas H, Kolodner RD. The Saccharomyces cerevisiae MLH3 gene functions in MSH3-dependent suppression of frameshift mutations. Proc Natl Acad Sci U S A 1998; 95:12404–9.

22 Jaafar L, Flores-Rozas H. Elucidating the role of human mismatch repair factor hMLH3. Cancer Biol Ther 2009;8:1421–3.

23 Hienonen T, Laiho P, Salovaara R, Mecklin JP, Järvinen H, Sistonen P, et al. Little evidence for involvement of MLH3 in colorectal cancer predisposition. Int J Cancer 2003;106:292–6.

24 Duraturo F, Liccardo R, Cavallo A, De Rosa M, Grosso M, Izzo P. Association of low-risk MSH3 and MSH2 variant alleles with Lynch syndrome: probability of synergistic effects. Int J Cancer 2011;129:1643–50.

25 Wu Y, Berends MJ, Sijmons RH, Mensink RG, Verlind E, Kooi KA, et al. A role for MLH3 in hereditary nonpolyposis colorectal cancer. Nat Genet 2001;29:137–8.

26 Liu HX, Zhou XL, Liu T, Werelius B, Lindmark G, Dahl N, et al. The role of hMLH3 in familial colorectal cancer. Cancer Res 2003; 63:1894–9.

27 Liccardo R, Lambiase M, Nolano A, De Rosa M, Izzo P, Duraturo F. Significance of rare variants in genes involved in the pathogenesis of Lynch syndrome. Int J Mol Med 2022;49.

28 Tsoulos N, Agiannitopoulos K, Potska K, Katseli A, Ntogka C, Pepe G, et al. The Clinical and Genetic Landscape of Hereditary Cancer: Experience from a Single Clinical Diagnostic Laboratory. Cancer Genomics Proteomics 2024;21: 448–63.

29 Salo-Mullen EE, Maio A, Mukherjee S, Bandlamudi C, Shia J, Kemel Y, et al. Prevalence and Characterization of Biallelic and Monoallelic NTHL1 and MSH3 Variant Carriers From a Pan-Cancer Patient Population. JCO Precis Oncol 2021; 5.

30 Mahmood K, Thomas M, Qu C, Hsu L, Buchanan DD, Peters U. Elucidating the Risk of Colorectal Cancer for Variants in Hereditary Colorectal Cancer Genes. Gastroenterology 2023;165:1070–6.e3.

31 Reijns MAM, Parry DA, Williams TC, Nadeu F, Hindshaw RL, Rios Szwed DO, et al. Signatures of TOP1 transcription-associated mutagenesis in cancer and germline. Nature 2022; 602:623–31.

32 Wainschtein P, Zhang Y, Schwartzentruber J, Kassam I, Sidorenko J, Fiziev PP, et al. Estimation and mapping of the missing heritability of human phenotypes. Nature 2025.

33. Valle L, Vilar E, Tavtigian SV, Stoffel EM. Genetic predisposition to colorectal cancer: syndromes, genes, classification of genetic variants and implications for precision medicine. J Pathol 2019;247:574–88.

34 Ten Broeke SW, van der Klift HM, Tops CMJ, Aretz S, Bernstein I, Buchanan DD, et al. Cancer Risks for PMS2-Associated Lynch Syndrome. J Clin Oncol 2018;36:2961–8.

35 Peltomäki P, Nyström M, Mecklin JP, Seppälä TT. Lynch Syndrome Genetics and Clinical Implications. Gastroenterology 2023;164:783–99.

36 Bjørnland T, Bye A, Ryeng E, Wisløff U, Langaas M. Powerful extreme phenotype sampling designs and score tests for genetic association studies. Stat Med 2018;37:4234–51.

37 Mefford J, Witte JS. The Covariate’s Dilemma. PLoS Genet 2012;8:e1003096.

38 Vali-Pour M, Park S, Espinosa-Carrasco J, Ortiz-Martínez D, Lehner B, Supek F. The impact of rare germline variants on human somatic mutation processes. Nat Commun 2022;13:3724.

39 Karczewski KJ, Francioli LC, Tiao G, Cummings BB, Alföldi J, Wang Q, et al. The mutational constraint spectrum quantified from variation in 141,456 humans. Nature 2020;581:434–43.

40 Benson G. Tandem repeats finder: a program to analyze DNA sequences. Nucleic acids research 1999;27: 573–80.

41 Jamal-Hanjani M, Hackshaw A, Ngai Y, Shaw J, Dive C, Quezada S, et al. Tracking genomic cancer evolution for precision medicine: the lung TRACERx study. PLoS biology 2014;12:e1001906.

42 Islam SMA, Díaz-Gay M, Wu Y, Barnes M, Vangara R, Bergstrom EN, et al. Uncovering novel mutational signatures by de novo extraction with SigProfilerExtractor. Cell Genom 2022;2:None.

43 Nunes L, Li F, Wu M, Luo T, Hammarström K, Torell E, et al. Prognostic genome and transcriptome signatures in colorectal cancers. Nature 2024;633:137–46.

