## Supplementary Figures for "Heterozygous germline *MSH3* mutations, and probably *MLH3* mutations, act as classical tumour suppressors, leading to excess somatic deletion mutations, signature ID4 and increased colorectal cancer risk"

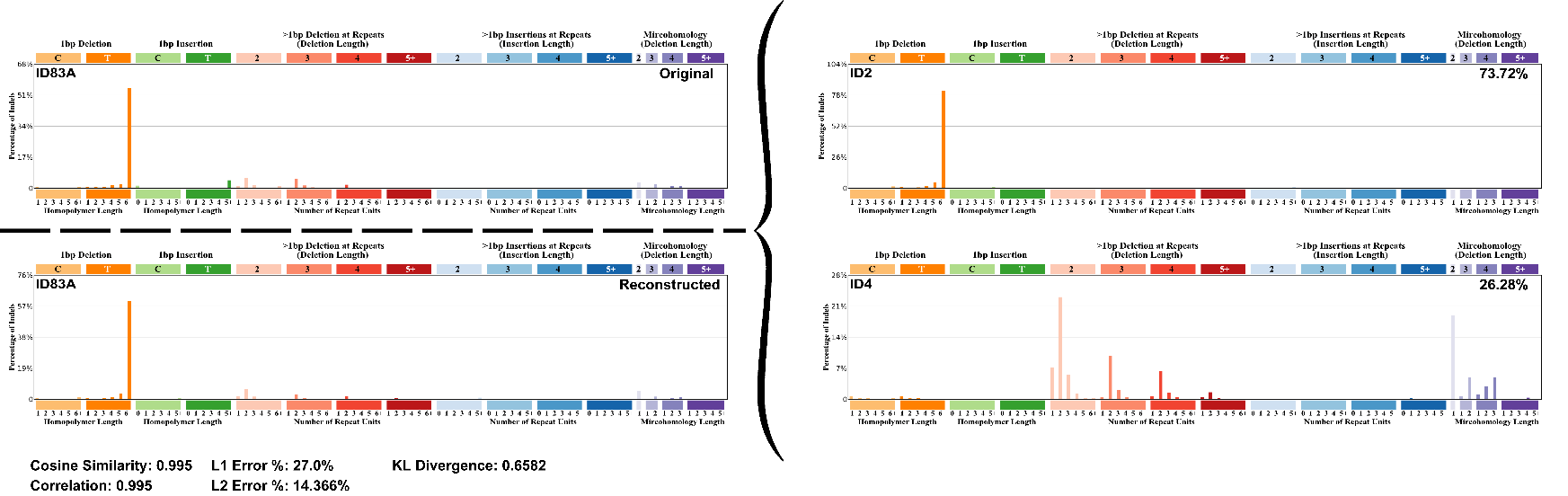

*Supplementary Figure 1.* *Decomposition of ID signatures ID83A, ID83B and ID83C extracted de novo from MSH3/MLH3-deficient cancers into COSMIC reference signatures.* In each of the three panels, the 83-channel plot in the top left quadrant shows one of the three signatures extracted *de novo* signature by SigProfileExtractor, termed ID83A, ID83B and ID83C respectively. This is then fitted to the COSMIC v3.2 reference set of ID signatures and the fitted signatures are shown in the right-hand quadrants of each panel. ID83A is fitted to a combination of ID2 and ID4, ID83B to ID4 alone and ID83C to ID1 and ID2 combined. The fitted signatures are then combined according to an assessed optimal weighting into a single signature decomposition output in the bottom left quadrant for each of ID83A, ID83B and ID83C. Goodness of fit metrics, such as cosine similarity, are provided for each of the decomposition outputs relative to its original *de novo* signature. In this case, the fit between each decomposed signature and its progenitor *de novo* signature is acceptable, such that the *de novo* signature can be regarded as being explained by one or more COSMIC reference signatures. A very similar process was followed when assessing CRCs individually, again resulting in ID4 detection in five or the six MSH3/MLH3-deficient CRCs.

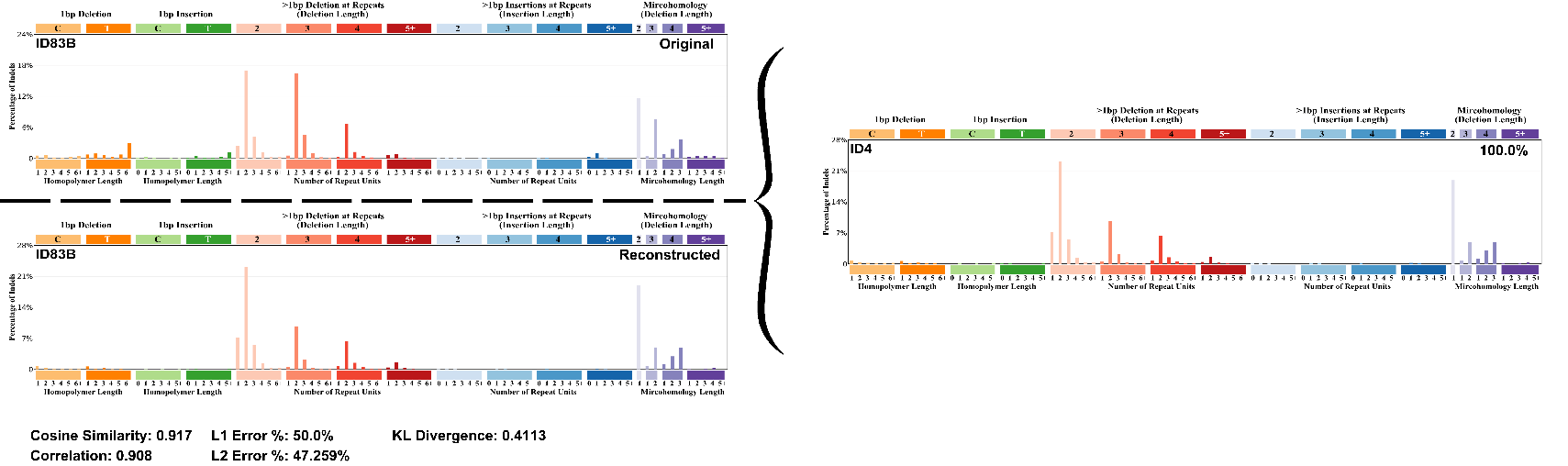

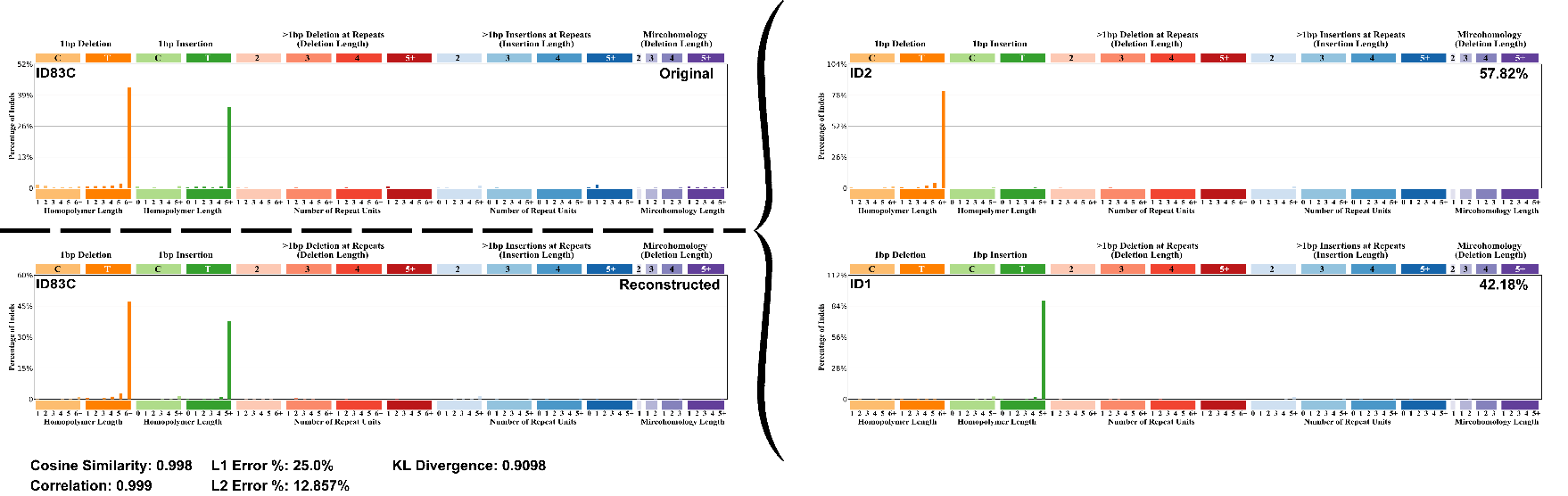

*Supplementary Figure 2.* *All de novo-extracted ID signatures in individual MSH3/MLH3-deficient cancers (six CRCs, two breast cancers).* Note that CRC-B11 was ID4-negative, although it did have an excess of mutations in the active ID4 channels compared with the bulk of MSS CRCs. The similarity of each of the other individual cancers to ID4 was high (cosine >0.90) when considering only the ≥2bp indel channels. ID1 and ID2 were the only other ID signatures present in these tumours.

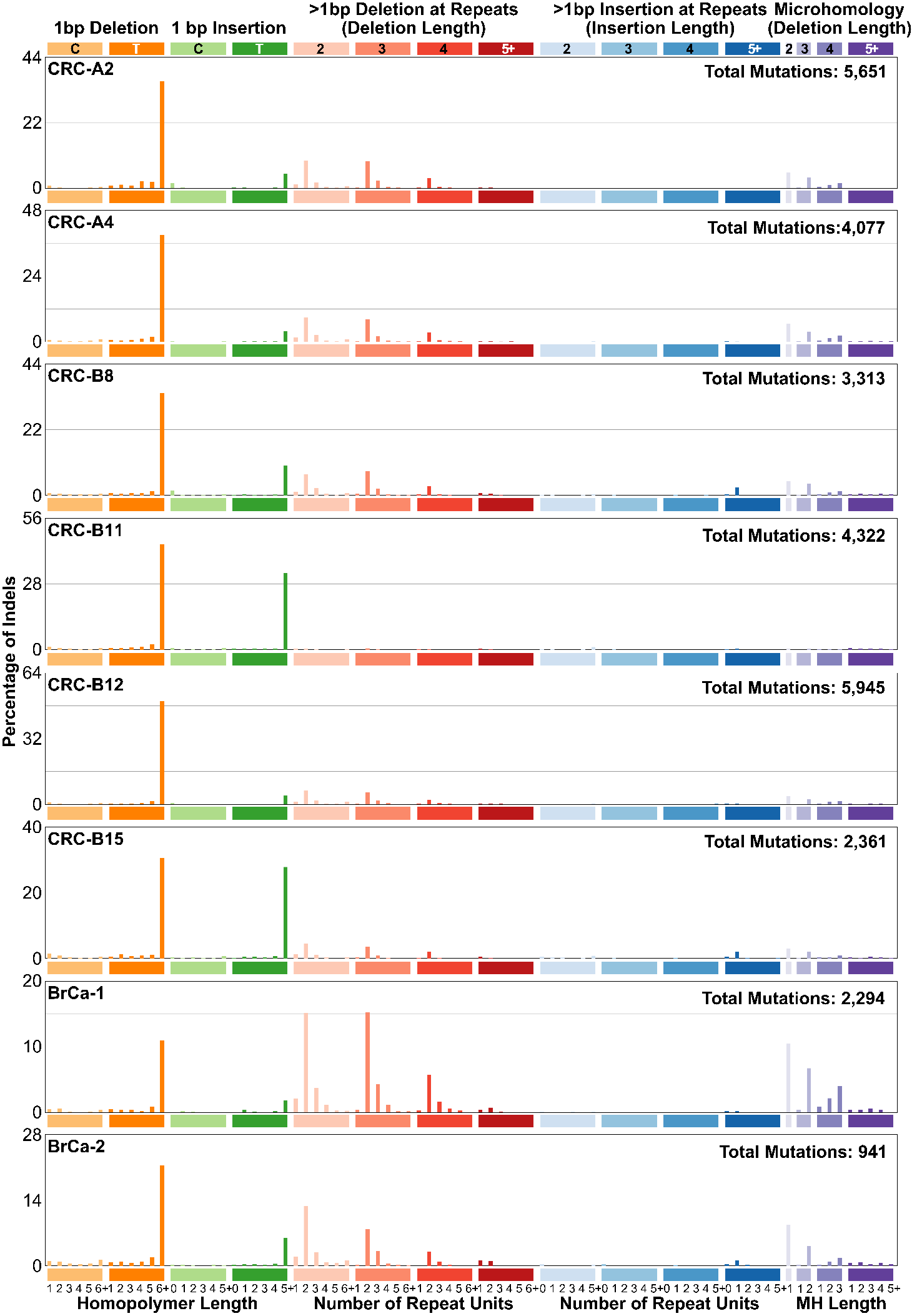

*Supplementary Figure 3. ID mutation spectrum resulting from subtraction for each channel of mean MSS bulk channel burden (wildtype for POLE, POLD1, MSH3 and MLH3) from mean MSH3/MLH3-deficient burden.* In experimental systems, such as mice and cells, it is possible to perform a subtraction-based analysis of mutation spectra, without extracting signatures. For example, if test animals carry a transgene expected to raise the mutation burden, its effects Even here, confounders could be present, *e.g.* if the increased mutation rate feeds through into higher proliferation or cell death. With caution, similar methodology can be applied to our cancers, since the underlying assumption – that MSH3/MLH3 deficiency simply adds mutations to those found in a typical MSS cancer – is consistent with the finding that in general, no mutational burdens – including specific ID channels or total SBSs, SVs and CNAs – are higher in bulk MSS than MSH3/MLH3-deficient cancers, suggesting that the former have not acquired features to ‘compensate’ for their MSH3/MLH3 proficiency (**Figure 3; Table 3; Supplementary Tables 1, 2 & 3**). (In fact, for 14 indel channels, counts were higher in the MSS bulk cancers, but all of these channels had low burdens, to the extent that the activities are not even visible on the plot below.) The plot below represents subtraction of bulk MSS ID mutation spectrum from the MSH3/MLH3-deficient CRC ID spectrum, with ID4 also shown for comparison. For the subtracted spectrum, cosine similarity to ID4 is high for ≥2bp indels (0.972), but much lower overall (0.424) owing to the 1:del:T:6+ excess in the MSH3/MLH3-deficient cancers that are, in effect, subsumed by ID2 when signatures are extracted (**Supplementary Figure 1**).

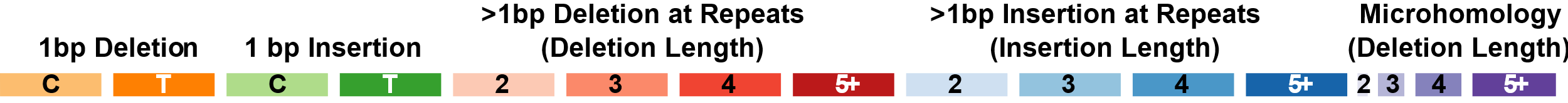

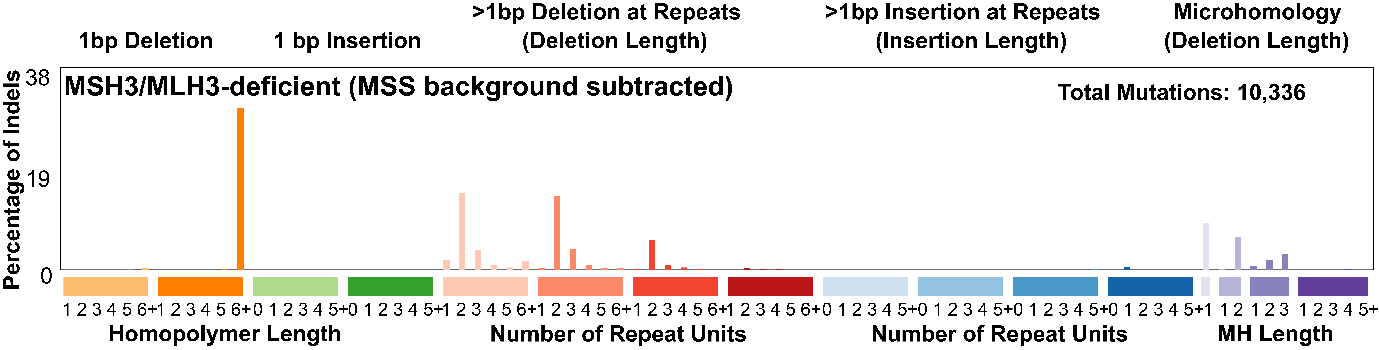

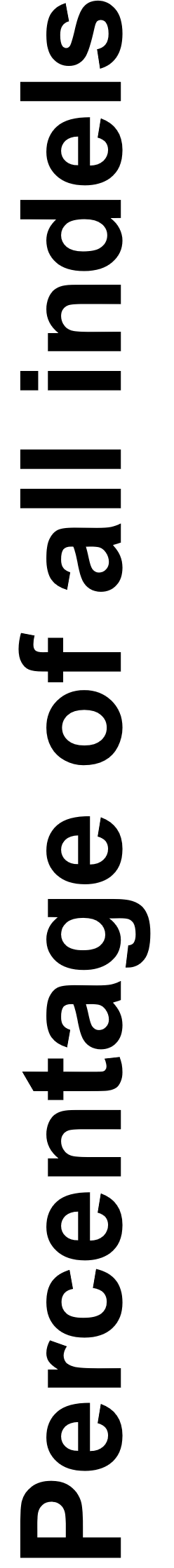

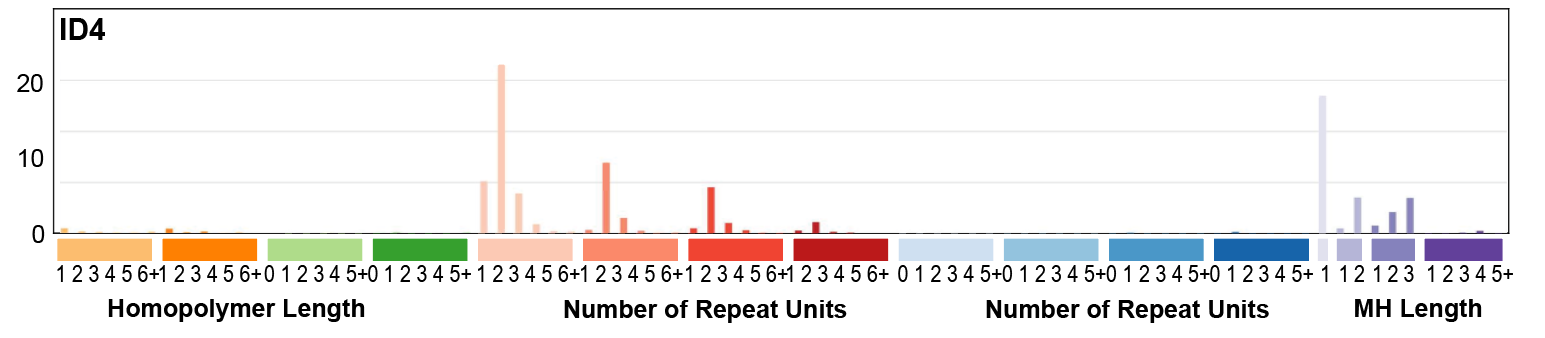

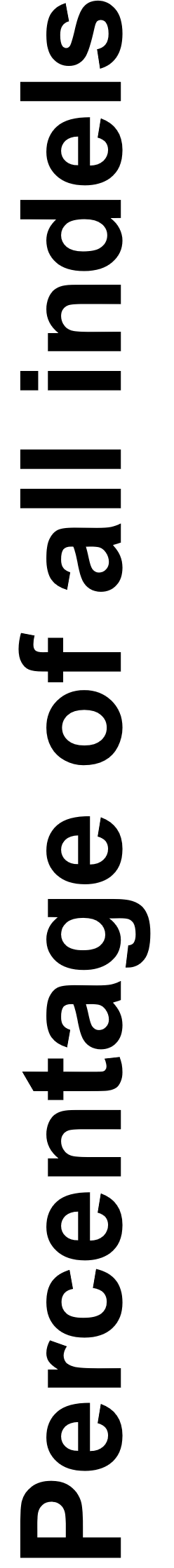

*Supplementary Figure 4. ≥2bp indel mutation spectra corresponding to (top to bottom) Reijns et al Rnaseh2b;Tp53 null mice, MSH3/MLH3-deficient CRCs from this study and ID4*. Bar heights are scaled to the most active channel for ease of comparison. There are several apparent differences, some of which may be inter-specific, but the clearest of these is the lower level of deletions at longer repeats in the mice relative to the CRCs, with ID4 intermediate between the other two data types. Cosine similarity between *Rnaseh2b;Tp53* null mice and ID4 is 0.866, compared with 0.913 for MSH3/MLH3-deficient CRCs and ID4. Cosine similarity between the mice and CRCs is quite modest at 0.668.

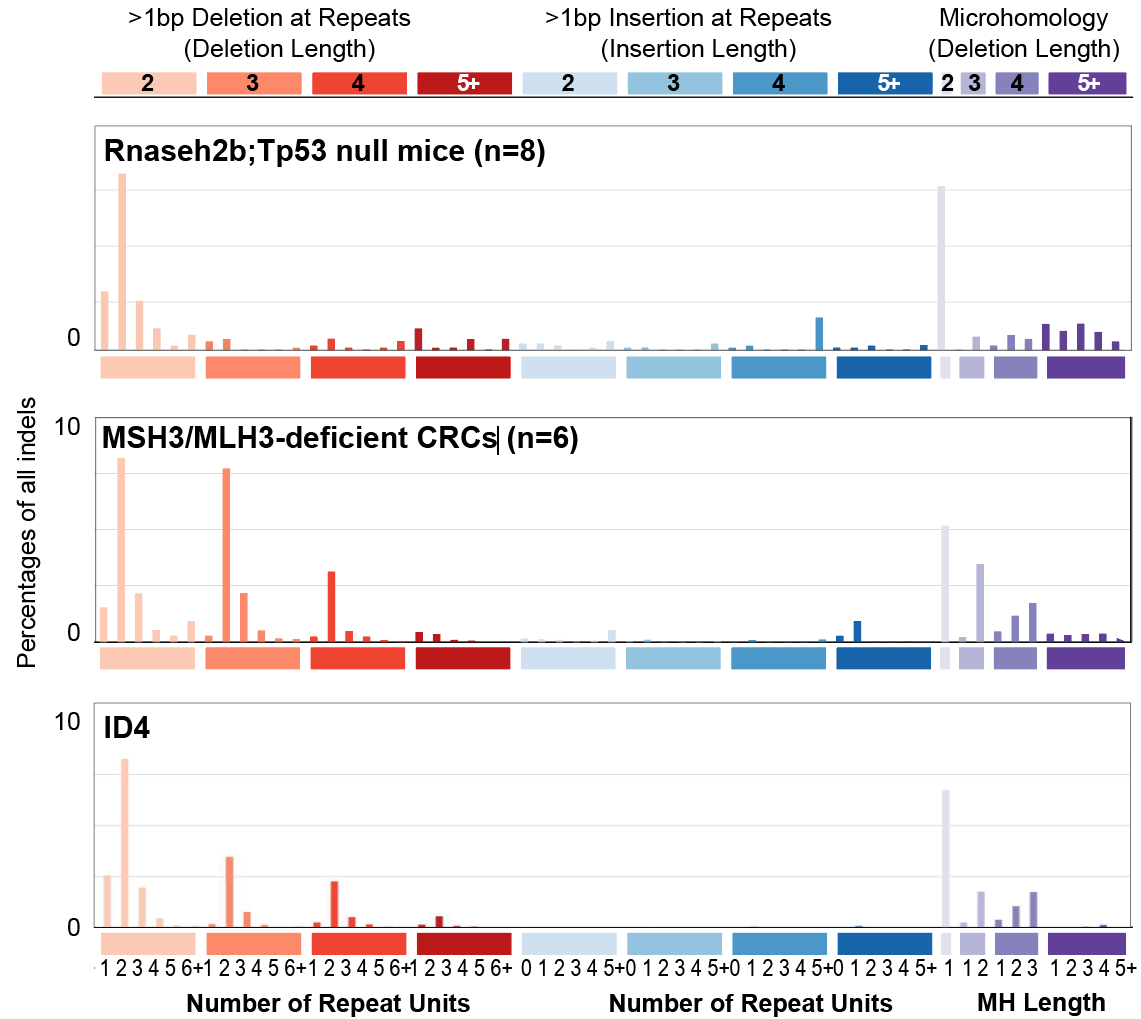

**Supplementary Tables**

1. CRC cases with pathogenic germline MSH3 or MLH3 variants from 100kGP: clinical and somatic mutation data are shown.

2. Germline pathogenic MSH3 and MLH3 variants in cases from 100kGP and CORGI, UK Biobank and Sweden, together with clinical and somatic mutation data where available.

3. The burdens and activities of insertion-deletion mutations in colorectal cancers according to MSH3/MLH3 and MSI+ status

4. Previous association studies with data on *MSH3* and *MLH3* heterozygotes.

5. ID4 presence/absence in extra-colonic cancers from carriers of germline heterozygous MSH3 or MLH3 mutations in 100kGP and second hits inactivating the germline wildtype allele.

6. Indel somatic mutation burdens and activities in breast cancers from 100kGP.
